# Low mitochondrial copy number drives atherogenic cardiovascular disease: evidence from prospective cohort analyses in the UK Biobank combined with Mendelian Randomization

**DOI:** 10.1101/2021.07.01.21259854

**Authors:** Jiao Luo, Raymond Noordam, J Wouter Jukema, Ko Willems van Dijk, Sara Hägg, Felix Grassmann, Saskia le Cessie, Diana van Heemst

## Abstract

**Background:** Mitochondrial DNA (mtDNA) content might be involved in the risk of cardiovascular disease. We aimed to investigate the association of mtDNA copy number (mtDNA-CN), as a proxy of mtDNA content, and coronary artery disease (CAD) and heart failure (HF) using multivariable adjusted and Mendelian Randomizations (MR) analyses.

**Methods:** Multivariable-adjusted analyses were conducted using Cox-proportional hazard models in 273,619 unrelated European descendants from UK Biobank (UKB). MtDNA-CN in peripheral blood cells was computed based on the weighted intensities of the mitochondrial genome probes. For the two-sample MR analyses, single nucleotide polymorphisms (SNPs) associated with mtDNA-CN were retrieved from genome-wide association studies in UKB. SNP-outcome associations were obtained for CAD from CARDIoGRAMplusC4D, UKB and FinnGen, comprising 902,538 participants (134,759 cases), and for HF from the HERMES consortium and FinnGen, collectively having data on 1,195,531 participants (70,706 cases). MR analyses were performed per database and results were subsequently meta analyzed using fixed-effects models per study.

**Results:** During a median follow-up of 11.8 years, participants in the lowest quintile of mtDNA-CN had higher risk for CAD (hazard ratio [95% CI]: 1.08 [1.03, 1.14]) and HF (hazard ratio [95% CI]: 1.15 [1.05, 1.24]) compared to those in the highest quintile. In MR analyses, the pooled odds ratios of genetically predicted per one-SD decrease in mtDNA were 1.16 (95% CI: 1.05, 1.27) for CAD and 1.00 (95% CI: 0.90, 1.10) for HF, respectively.

**Conclusion:** Our findings support a possible causal role of lower mtDNA-CN in higher CAD risk, but not in higher HF risk.

## 1. Introduction

Cardiovascular diseases (CVD) are the leading cause of death worldwide. The role of mitochondria in CVD has received growing attention due to the high energy demands of cardiomyocytes. Mitochondrial dysfunction, considered a hallmark of the aging process (1), leads to reduced bioenergetic capacity and disrupted redox homeostasis, and is hypothesized to be an important component in the pathogenesis of CVD (2,3). In addition to nuclear genes, the mitochondrial DNA (mtDNA), which constitutes the mitochondria’s own genome, plays a key role in energy generation during oxidative phosphorylation, functioning to encode 27 genes that are essential for the synthesis of multi-subunit enzymatic components of the electron transport chain. Although mitochondria contain a variable number of mtDNA copies, mitochondrial DNA copy number (mtDNA-CN) is considered to approximate the average number of mitochondria per cell and reflects the activity of mitochondrial enzymes and their bioenergetic capacity, thus serving as a surrogate biomarker of mitochondrial function.

Recent epidemiological studies have assessed the associations between peripheral blood mtDNA-CN and multiple cardiovascular endpoints. These studies so far unequivocally indicated lower mtDNA-CN as an independent risk factor of prevalent CVD in case-control and retrospective cohort studies (4-7), and of incident cardiovascular disease and risk of sudden cardiac death in the prospective Atherosclerosis Risk in Communities (ARIC) study (8-11). Nevertheless, apart from the ARIC study, other studies comprised a small sample size and/or limited number of cases, which might have resulted in insufficient statistical power, undermining the reliability. In addition, potential confounding factors were not fully taken into account in previous studies, such as sleep or physical activity, which are possibly related to both mtDNA-CN and CVD. Furthermore, due to the vague onset and long-term progression of CVD pathogenesis, it is not possible to fully eliminate reverse causation in studies with observational study designs.

Triangulation of causal inference in etiological epidemiology was described previously by Lawlor *et al*., which integrates results from different methodological approaches to enhance the reliability of a research study (12). The confidence of the findings will be strengthened if results from different approaches are consistent. Based on earlier studies, we hypothesized that lower mtDNA-CN is associated with an increased risk of incident CVD. In the present study, we aimed to firstly prospectively examine the associations between mtDNA-CN, and incidence of coronary artery disease (CAD) and heart failure (HF) in participants of European ancestry in the UK Biobank (UKB), taking several possible measured confounding factors into account. In addition, Mendelian Randomization (MR) uses genetic instruments associated with exposures of interest to infer causal associations between life-long risk factors (exposure) on diseases (outcome) in the absence of reverse causation and residual confounding (13). Therefore, we additionally conducted MR analyses to examine whether the associations (if any) observed between mtDNA-CN and CAD and HF from prospective studies may be causal.

## 2. Methods

### Prospective study

#### Study population

The UKB cohort is a prospective cohort with 502,628 participants between the age of 40 and 69 years recruited from the general population at multiple assessment centers across the UK between 2006 and 2010. Invitation letters were sent to eligible adults registered to the National Health Services (NHS) and living within a 25 miles distance from one of the assessment centers. Participants provided information on their lifestyle and medical history through touch-screen questionnaires and physical measurements. Blood samples were collected for genotyping. The UKB study was approved by the North-West Multi-center Research Ethics Committee (MREC). Access for information to invite participants was approved by the Patient Information Advisory Group (PIAG) from England and Wales. All participants provided electronic written informed consent for the study. Detailed information about the study design, investigation methods, ethical approval as well as limitations have been reported previously (14).

Full genetic data release (July 2017) that included genotype data from 488,377 individuals were used in the present study. Further details of the array design, genotyping and imputation procedures have been described elsewhere (15). Participants who did not pass the sample quality control were initially excluded according to the parameters presented in the sample quality control file of the UKB, including participants who were: 1) not used to compute principal components; 2) identified as outliers in heterozygosity and missing rates, which is indicative of poor-quality genotype data for these samples; 3) identified as putatively sex chromosome aneuploidy; 4) sex inferred from genotype data did not match their self-reported sex; 5) have an excessive number (more than 10) of relatives in the database. To minimize the variation resulted from population substructures, we restricted the study population to unrelated white British individuals. Participant relatedness was available in the UKB by estimating kinship coefficients for all pairs. White British ancestry was identified as self-reported ethnic background, and further refined the population definition in a principal component analysis of the genotype data that were tightly clustered. This resulted in a primary study cohort comprising 302,685 unrelated European ancestry participants. A flowchart on the different exclusions is provided in **Figure S1**.

#### mtDNA-CN computation

The exposure of interest for our analysis is the somatic mtDNA-CN as assessed from the intensities of genotyping probes on the mitochondrial chromosome on the Affymetrix Array. The method for computing mtDNA-CN has been described in detail previously (16). We followed the same analysis pipeline to calculate mtDNA-CN in the available data of UKB (https://github.com/GrassmannLab/MT_UKB). In brief, the relative amount of mtDNA hybridized to the array at each probe was log2 transformed ratio (L2R) of the observed genotyping probe intensity divided by the intensity at the same probe observed in a set of reference samples. The median L2R values across all 265 variants passing quality control on the MT chromosome were used as an initial raw measure of mtDNA-CN. To correct for the confounding induced by poorly performing probes, we weighted the L2R values of each probe multiplied by the weight of the probe that were generated from a multivariate linear regression model in which those intensities statistically significantly predicted normalized mitochondrial coverage from exome sequencing data, resulting in a single mtDNA-CN estimate for each individual. To eliminate the plate effect, we subsequently normalized the CN to a mean of zero and standard deviation (SD) of one within each genotyping plate comprising 96 wells. An additional quality control step was performed by eliminating individuals with high standard deviation (SD) (two SD from the mean) of autosomal probes log2 ratio (L2R). Consequently, 293,245 individuals remained in the cohort.

#### Outcome definition

Outcomes in the analysis were incident cardiovascular diseases during the time period from recruitment to January 1^st^, 2021. Incident disease status was ascertained by linkage with hospital admissions data and national death register data to identify the date of the first known CVD or CVD-related death after the date of baseline assessment. The linkage details are presented in the original study protocol (https://www.ukbiobank.ac.uk/media/gnkeyh2q/study-rationale.pdf, accessed April 2021). The outcomes were incident CAD and HF, separately. Incident disease diagnoses are coded according to the International Classification of Diseases edition 10 (ICD-10); CAD cases were defined as angina pectoris (I20), myocardial infarction (MI) (I21 and I22), and acute and chronic ischemic heart disease (IHD) (I24 and I25); Incident HF cases were defined as I50. In addition, we analyzed acute myocardial infarction (MI) and chronic ischemic heart disease (IHD) as separate outcomes in sensitivity analyses. Follow-up time is computed from the baseline visit to the diagnosis of incident disease, death, or the censoring, whichever occurred first.

#### Covariates definition

Covariates used in the study were from baseline measurements, which included demographic parameters (age at recruitment, sex, deprivation index); the first and second principal components (PCs) to correct for possible remaining population stratification (17); genotyping batch; cell numbers (white blood cell counts and platelet counts); anthropometric measure of body mass index (BMI) in kg/m^2^; self-reported lifestyle factors (smoking status [never, past and current], alcohol consumption frequency [twice or less per week/ more than three times per week], physical activity [MET hours per week for moderate-vigorous activity], sleep duration in hours and insomnia symptoms [yes/no]); familial CVD history (yes/no), lipid levels (mmol/l) (total and LDL [low-density lipoprotein] cholesterol) lipid-lowering medication, blood pressure (mmHg, average of the two measurements when applicable) and blood pressure-lowering medication, as well as baseline type 2 diabetes mellitus (T2DM, yes/no) from the medical records.

#### Statistical analysis

After further exclusion of participants with any prevalent cardiovascular disease (including CAD, HF and stroke) or withdrawn informed consent, the study cohort comprised an analytic sample of 273,619 individuals (**Figure S1**). Baseline characteristics of the study population were described in quintiles of mtDNA-CN and presented as mean (SD) or median (interquartile range, IQR) for continuous variables and frequency (proportion) for categoric variables. Cumulative incidence for competing risks (CICR) was used to plot the cumulative incidence of both CAD and HF against follow-up time by mtDNA-CN quintiles, accounting for death as a competing event.

Cox proportional hazards models were used to estimate hazard ratios (HR) and corresponding 95% confidence intervals (CI) for the association between mtDNA-CN and incident CAD and HF separately. Two multivariable-adjusted regression models were fitted: Model 1 was adjusted for age, sex, the first and second PCs, genotyping batch, white blood cell count and platelet count; Model 2 was additionally adjusted for BMI, smoking, alcohol consumption, sleep duration, insomnia, physical activity, familial CVD history, lipid levels and lipid-lowering medication, blood pressure and blood pressure-lowering medication and T2DM. In the primary analysis, we treated mtDNA-CN as a continuous variable and assessed the risk of incident diseases associated with per one-SD decrease in mtDNA-CN. Subsequently, mtDNA-CN was categorized into quintiles, and hazard ratios (HR) compared the 1st to 4th quintiles with the 5th quintile (reference). The proportional hazard assumption was graphically assessed by plotting log(-log[survival]) versus log(follow-up time), and was tested using Schoenfeld residuals. As secondary analysis, to test the possible nonlinear dose-response associations between mtDNA-CN and incident diseases, we implemented restricted cubic splines with knots located at 5th, 50th and 95th percentiles.

Missing data were present in the covariates, and these were subsequently imputed using multiple imputation by chain equations (MICE) (18), setting the number of imputed datasets to 10. We used predictive mean matching for continuous variables, logistic regression for binary variables and polytomous regression for categorical variables. The imputation model included mtDNA-CN, all covariates, the Nelson-Aalen estimator of cumulative hazard and incident disease status. Cox proportional hazards models were fitted within each imputed dataset and were subsequently pooled according to Rubin’s rules.

As sensitivity analysis, firstly, interaction terms between mtDNA-CN and age, sex were added to Model 2 to test for the presence of effect modification by sex or age, and subgroup analyses were also performed in each stratum of sex and age (< 50 years, 50∼60 years, > 60 years), respectively. Secondly, all analyses were performed for the CAD subtypes, i.e. MI and IHD. Thirdly, analyses were repeated restricting to participants without missing data on covariates, i.e. complete cases (N = 162,002)

### Mendelian Randomization (MR)

#### Study design

In addition to the multivariable adjusted regression analyses, we also leveraged both two-sample and one-sample MR (for UKB analyses) to examine whether there is evidence for possible causality. Three main instrumental variable assumptions are usually invoked, which include that instruments must be 1) associated with exposure, 2) independent of any confounding factors between exposure and outcome, and 3) independent of outcome conditioning on exposure and all confounding factors between exposure and outcome.

#### Instrumental variables

We retrieved 129 independent SNPs (linkage disequilibrium < 0.05) nuclear single-nucleotide polymorphisms (SNPs) as genetic instruments that were associated with continuous mtDNA-CN at a genome-wide significance threshold (p < 5e-08), as identified in a recent genome-wide association study (GWAS) by Longchamps *et al* (19). The study was performed in 465,809 individuals of White European ancestry combining the Cohorts for Heart and Aging Research in Genomic Epidemiology (CHARGE) consortium and UKB. Genetic associations were adjusted for age, sex and covariates that were specific in each cohort, such as PCs, blood collection sites, family structure and cell composition. F statistics [(β/se)^2^] were computed to evaluate instrumental strength, and SNPs with a value of less than 10 was considered as weak instruments. Furthermore, we calculated the proportion of total variance in the exposure explained by each instrument (R^2^) separately (20).

#### Outcome data source

Summary statistics for instrument-CAD associations were extracted from 3 large databases separately, the CARDIoGRAMplusC4D (Coronary Artery Disease Genome-Wide Replication and Meta-analysis plus the Coronary Artery Disease Genetics) consortium, UKB, and FinnGen study (freeze 5, released in May 2021). Similarly, summary statistics for SNP-HF associations were drawn from HERMES Consortium (Heart Failure Molecular Epidemiology for Therapeutic Targets Consortium, which included data from UKB in the meta-analysis) and the FinnGen study, respectively. The descriptions, number of cases and controls, cases definition as well as covariates used for associations tests of each of the databases are presented in detail in **Supplementary Text** and **Table S1**.

#### Mendelian Randomization analysis

SNP-exposure and SNP-outcome data were harmonized to make alignment on effect alleles. Palindromic SNPs were eliminated (21). The primary MR analysis was performed using Inverse-variance weighted (IVW) method to combine the SNP-specific estimates calculated using Wald ratios, assuming all instrumental variables are valid (22). Results were expressed as an odds ratio (OR) on disease risk for a one-SD decrease in genetically predicted mtDNA-CN. When the MR assumptions were met, this OR approximated the causal effect of the exposure on the outcome. Sensitivity analyses accounting for pleiotropy were conducted, including Weighted-Median Estimator (WME) and MR-Egger regression (23,24), both of which assume that at least half of the instrumental variable had to be valid. The intercept from MR-Egger represents the average pleiotropic effect; when the intercept deviates from zero, estimates from IVW might be biased. MR-PRESSO (MR Pleiotropy RESidual Sum and Outlier) was applied to detect and correct for horizontal pleiotropy through removing outliers (25). Moreover, we examined the heterogeneity using Cochran’s Q statistic among all SNPs within each outcome database.

For outcomes derived from the UKB, despite the gene-exposure associations were from the same population, it has been shown that Two-sample MR methods can be reliably used for one-sample MR performed within large biobanks, such as UKB, with the exception of the MR-Egger sensitivity analysis (26).

#### Meta-analysis of estimates from different databases

The effect of mtDNA-CN on CAD/HF in MR analyses were separately estimated in different outcome databases separately, CARDIoGRAMplusC4D consortium (CAD) or HERMES (HF), UKB (CAD only) and FinnGen (both), and derived estimates were subsequently pooled using fixed-effects meta-analysis.

#### Sensitivity analysis

Despite the large sample size of the GWAS used for the selection of instrumental variables in Longchamps *et al* study, which increased the statistical power, the assessments of mtDNA-CN among cohorts that contributed data to the meta-analysis were very different. To account for this heterogeneity, we additionally performed sensitivity analyses restricting to genetic instruments identified in the UKB only. Therefore, 66 independent (linkage disequilibrium < 0.1) SNPs were used that were associated with mtDNA-CN at a genome-wide significance threshold (p < 5e-08) from 295,150 participants conducted by Hägg *et al*. (16). Genetic associations were adjusted for PCs, age, sex, genotyping batch, genotyping missingness/call rate and cell composition. All MR analyses were repeated with the substitution of the 69 genetic instruments for mtDNA-CN.

All the analyses were performed using R (v3.6.3) statistical software (The R Foundation for Statistical Computing, Vienna, Austria). Packages used in the analyses included “cmprsk” for cumulative incidence for competing risk analyses, “mice” for multiple imputations, “survival” and “survminer” for cox proportional hazard regression, “rms” for nonlinear dose-response associations, “TwoSampleMR” for MR analyses (27), and “meta” for meta-analyses (28). All results were reported as odds ratio with accompanied 95% confidence interval.

## 3. Results

### Prospective results

#### Main analyses

A total of 273,619 participants were eligible for analyses after exclusion (**Figure S1**). Compared with the highest quintile of mtDNA-CN (**Table 1**), participants in the lower quintiles were more likely to have unfavorable CVD risk factors, including older age, male sex, higher BMI, higher blood pressure and more blood pressure-lowering medication, higher lipids (total cholesterol and LDL) and more cholesterol lowering medication, less physical activity, more current smokers and a higher percentage of familial history of CVD or prevalent T2DM.

**Table 1.** Baseline charactertistics of the study participants by quintiles of mtDNA copy number.

| Variable | Q1 | Q2 | Q3 | Q4 | Q5 |
| --- | --- | --- | --- | --- | --- |
| N | 54,724 | 54,724 | 54,723 | 54,724 | 54,724 |
| Age (years) | 57.2 (8.0) | 56.8 (8.0) | 56.5 (8.0) | 56.2 (8.0) | 55.9 (7.9) |
| Sex |  |  |  |  |  |
| Male | 25,413 (46.4%) | 24,898 (45.5%) | 24,478 (44.7%) | 24,320 (44.4%) | 23,622 (43.2%) |
| Female | 29,311 (53.6%) | 29,826 (54.5%) | 30,245 (55.3%) | 30,404 (55.6%) | 31,102 (56.8%) |
| BMI (kg/m<sup>2</sup>) | 27.6 (5.0) | 27.4 (4.8) | 27.3 (4.6) | 27.1 (4.6) | 26.9 (4.5) |
| Deprivation index | -1.5 (2.9) | -1.6 (2.9) | -1.7 (2.9) | -1.7 (2.9) | -1.7 (2.9) |
| Diastolic blood pressure (mmHg) | 82.9 (10.2) | 82.6 (10.0) | 82.4 (10.0) | 82.3 (10.1) | 81.8 (10.1) |
| Systolic blood pressure (mmHg) | 139.5 (18.9) | 138.7 (18.7) | 138.2 (18.7) | 137.6 (18.4) | 136.6 (18.3) |
| Blood pressure-lowering medication |  |  |  |  |  |
| Yes | 10,826 (19.8%) | 10,085 (18.4%) | 9598 (17.5%) | 9089 (16.6%) | 8524 (15.6%) |
| No | 43,898 (80.2%) | 44,637 (81.6%) | 45,128 (82.5%) | 45,635 (83.4%) | 46,199 (84.4%) |
| Total cholesterol (mmol/L) | 5.8 (1.1) | 5.8 (1.1) | 5.8 (1.1) | 5.8 (1.1) | 5.7 (1.1) |
| HDL (mmol/L) | 1.5 (0.4) | 1.5 (0.4) | 1.5 (0.4) | 1.5 (0.4) | 1.5 (0.4) |
| LDL (mmol/L) | 3.7 (0.9) | 3.6 (0.8) | 3.6 (0.9) | 3.6 (0.8) | 3.6 (0.8) |
| Triglycerides (mmol/L) | 1.8 (1.0) | 1.8 (1.0) | 1.7 (1.0) | 1.7 (1.0) | 1.7 (1.0) |
| Cholesterol lowering medication |  |  |  |  |  |
| Yes | 7596 (13.9%) | 7394 (13.5%) | 7148 (13.1%) | 6822 (12.5%) | 6663 (12.2%) |
| No | 47128 (86.1%) | 47330 (86.5%) | 47575 (86.9%) | 47902 (87.5%) | 48061 (87.8%) |
| Physical activity (moderate-vigorous MET hours/week) | 26.5 (33.8) | 27.0 (34.3) | 27.1 (34.6) | 27.1 (33.8) | 27.1 (33.8) |
| Alcohol consumption frequency |  |  |  |  |  |
| At least three times per week | 24,939 (45.6%) | 24,836 (45.4%) | 25,303 (46.2%) | 25,400 (46.4%) | 25,679 (46.9%) |
| Twice or less per week | 29,751 (54.4%) | 29,851 (54.5%) | 29,381 (53.7%) | 29,287 (53.5%) | 29,015 (53.0%) |
| Data missing | 34 (0.1%) | 37 (0.1%) | 39 (0.1%) | 37 (0.1%) | 30 (0.1%) |
| Smoking status |  |  |  |  |  |
| Current | 6124 (11.2%) | 5643 (10.3%) | 5369 (9.8) | 5046 (9.2%) | 4521 (8.3%) |
| Previous | 18,646 (34.1%) | 18831 (34.4%) | 18588 (34.0%) | 18586 (34.0%) | 18702 (34.2%) |
| Never | 29,732 (54.3%) | 30079 (55.0%) | 30594 (55.9%) | 30914 (56.5%) | 31350 (57.3%) |
| Data missing | 222 (0.4%) | 171 (0.3%) | 172 (0.3%) | 178 (0.3%) | 151 (0.3%) |
| Sleep duration (hours) | 7.1 (1.2) | 7.1 (1.2) | 7.1 (1.2) | 7.1 (1.2) | 7.1 (1.2) |
| Insomnia |  |  |  |  |  |
| Sometimes | 26,161 (47.8%) | 26,273 (48.0%) | 26,420 (48.3%) | 26,243 (48.0%) | 26,321 (48.1%) |
| Usually | 15,488 (28.3%) | 15,145 (27.7%) | 15,031 (27.5%) | 14,982 (27.4%) | 14,991 (27.4%) |
| Never/rarely | 13,038 (23.8%) | 13,271 (24.3%) | 13,237 (24.2%) | 13,454 (24.6%) | 13,384 (24.5%) |
| Data missing | 37 (0.1%) | 35 (0.1%) | 35 (0.1%) | 45 (0.1%) | 28 (0.1%) |
| Familial CVD history |  |  |  |  |  |
| Yes | 21,539 (39.4%) | 21,716 (39.7%) | 21,690 (39.6%) | 21,687 (39.6%) | 21,359 (39.0%) |
| No | 27,791 (50.8%) | 27,822 (50.8%) | 28,005 (51.2%) | 28,026 (51.2%) | 28,420 (51.9%) |
| Data missing | 5394 (9.8%) | 5186 (9.5%) | 5028 (9.2%) | 5011 (9.2%) | 4945 (9.0%) |
| T2DM history (yes) |  |  |  |  |  |
| Yes | 1184 (2.2%) | 1093 (2.0%) | 969 (1.8%) | 914 (1.7%) | 879 (1.6%) |
| No | 53,540 (97.8%) | 53,631 (98.0%) | 53,755 (98.2%) | 53,810 (98.3%) | 53,845 (98.4%) |
| Follow-up time |  |  |  |  |  |
| CAD | 11.6 (10.8, 12.5) | 11.7 (10.9, 12.5) | 11.8 (11.0, 12.5) | 11.8 (11.1, 12.5) | 11.9 (11.2, 12.5) |
| HF | 11.7 (10.9, 12.5) | 11.8 (11.0, 12.5) | 11.8 (11.1, 12.6) | 11.9 (11.2, 12.6) | 11.9 (11.3, 12.6) |

Data are mean (SD) or median (interquartile range, IQR) for continuous variables and frequency (percentage) for categorical variables. Some percentages do not add up to 100 because of rounding. BMI: Body mass index; HDL: high-density lipoprotein cholesterol; LDL: low-density lipoprotein cholesterol; CVD: cardiovascular disease; CAD: coronary artery disease; HF: heart failure.

During a median follow-up of 11.8 (interquartile range: 11.0, 12.5) years, 18,346 participants developed CAD and 5795 participants developed HF. Cumulative incidence of both CAD and HF increased stepwise with the decrease in mtDNA-CN, accounting for death as a competing risk (p for Gray’s test < 0.001) (**Figure 1**). In multivariable-adjusted cox proportional hazard models (model 1), a one-SD decrease in mtDNA-CN was associated with 1.06-fold (95% confidence interval, CI: 1.05, 1.08) and 1.09-fold (95%CI: 1.06, 1.12) higher hazard of CAD and HF, respectively; adjusted HRs for the first versus the fifth (reference) quintile of mtDNA-CN were 1.18 (95%CI: 1.23, 1.24) for CAD, and 1.28 (95%CI: 1.17, 1.39) for HF. Additional adjustment for CVD risk factors only minimally attenuated the estimates of CAD and HF (**Figure 2** and **Table S2**). Restricted cubic spline analyses confirmed an approximately linear dose-response relationship between lower mtDNA-CN with the higher risk of CAD (p for non-linearity = 0.14) and HF (p for non-linearity = 0.73), as shown in **Figure 3**.

**Figure 1.**
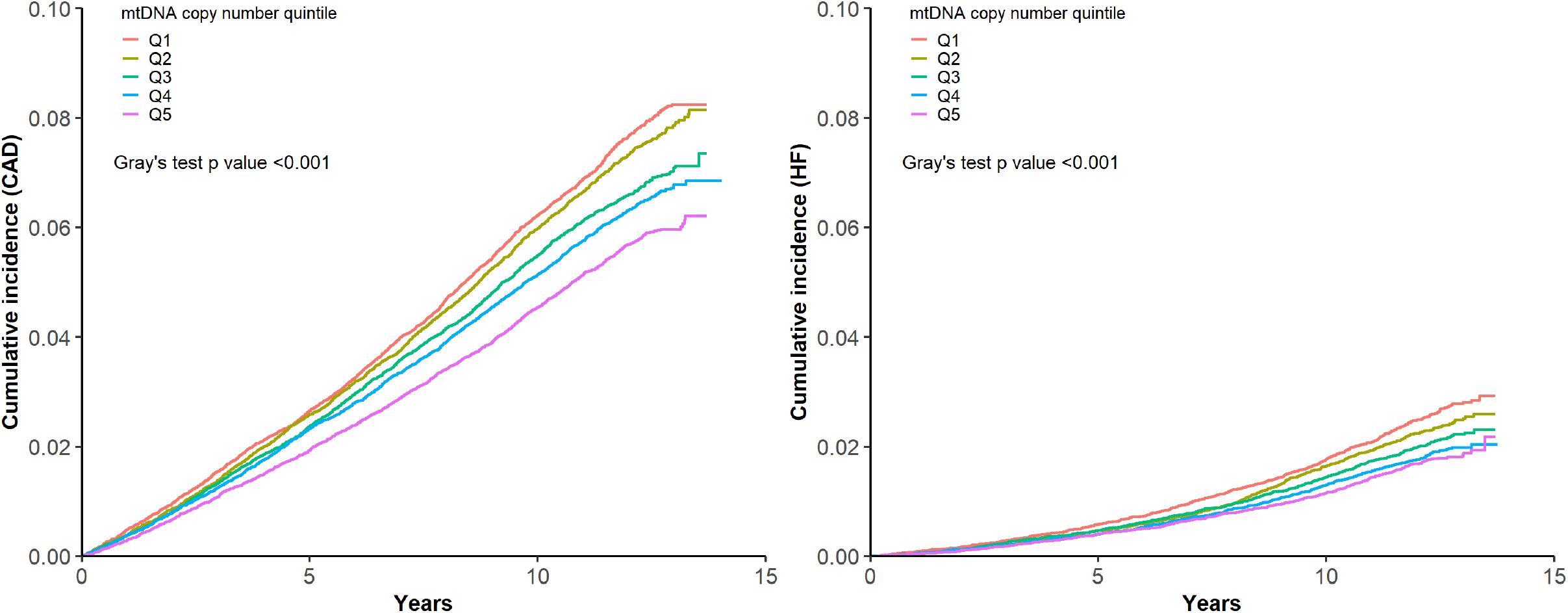
Cumulative Incidence of CAD and HF by quintiles of mtDNA copy number. We calculated Cumulative incidence for CAD and HF, accounting for death as a competing event. Differences in cumulative incidence between mtDNA copy number quintiles were assessed using Gray’s test. CAD: coronary artery disease; HF: heart failure.

**Figure 2.**
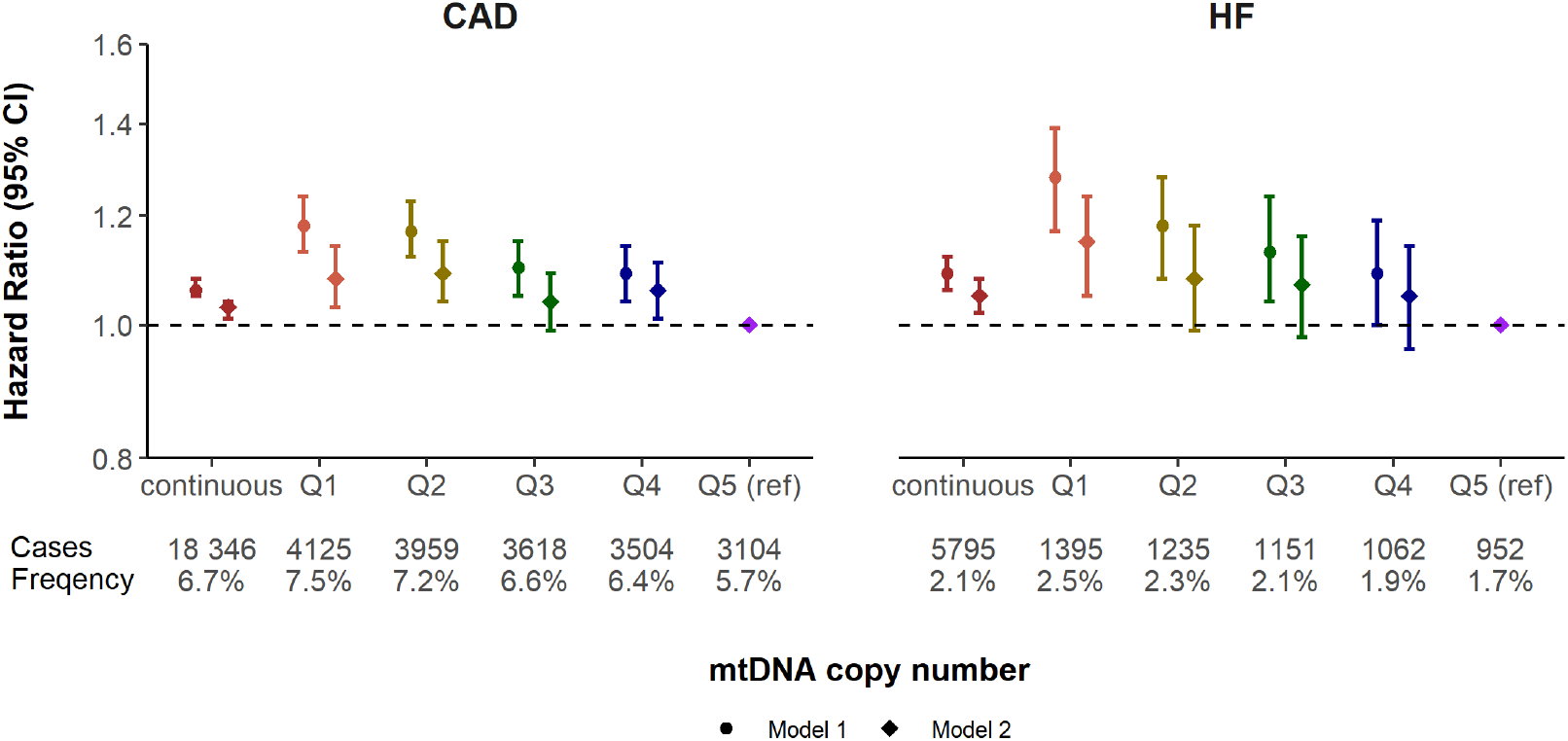
Hazard ratios for incident CAD and HF by quintiles of mtDNA copy number. Estimated hazard ratios for the effect of per-SD decrease in mtDNA copy number (continuous), or for the 1st to the 4th quintile compared to the 5th (reference) quintile (categorical) on CAD and HF. Model 1 was adjusted for age, sex, genotyping batch, the first two principal components, white blood cell count and platelet count. Model 2 was model 1 additionally adjusted for body mass index, physical activity, smoking status, alcohol consumption frequency, blood pressure and blood pressure-lowering medication, cholesterol, triglycerides and lipid-lowering medication, sleep duration and insomnia, type 2 diabetes status, and familial history of cardiovascular disease. CAD: coronary artery disease; HF: heart failure.

**Figure 3.**
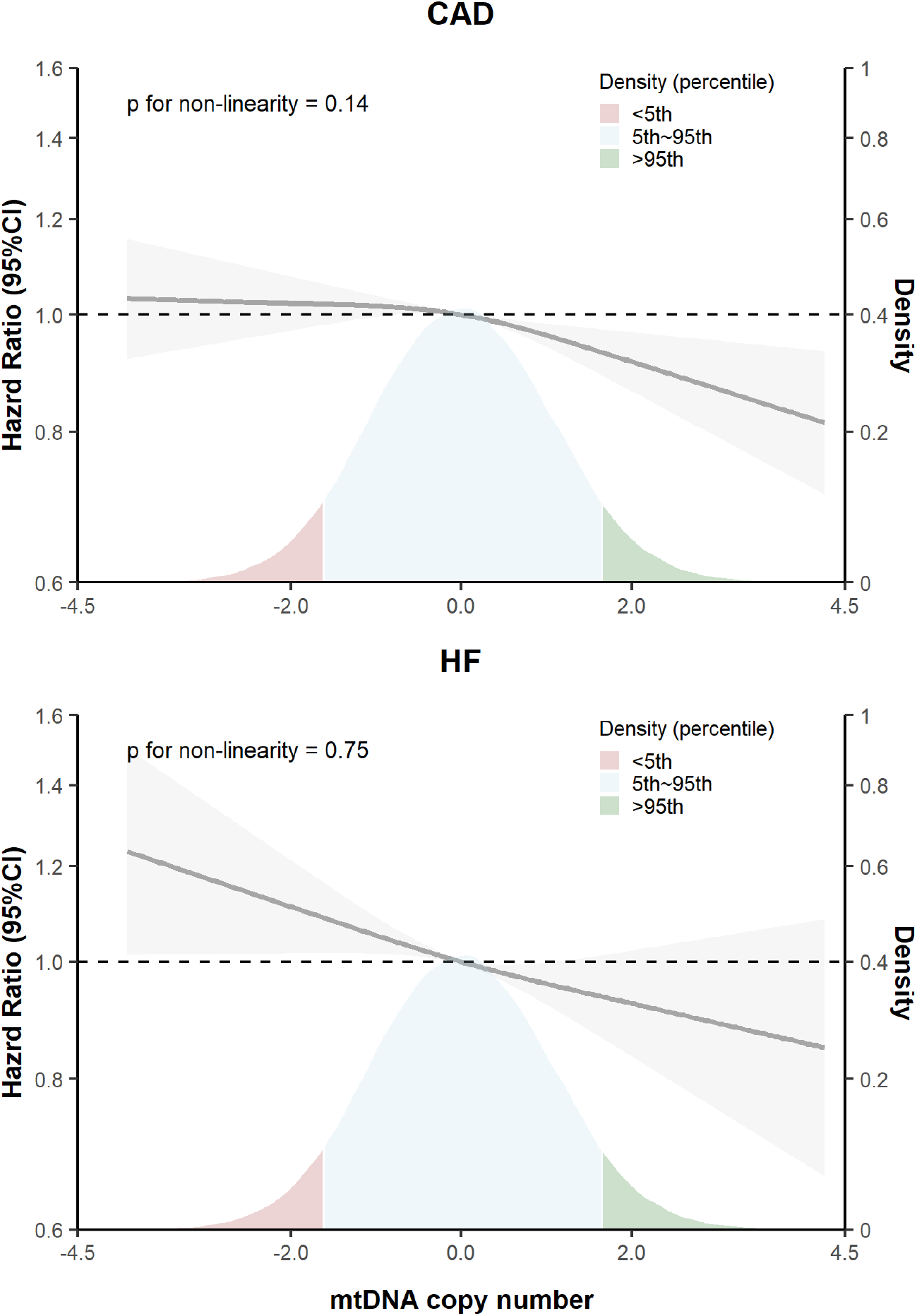
Hazard ratios for incident CAD and HF by levels of mtDNA copy number. Solid lines represented hazard ratios (derived from model 2 adjusted for age, sex, genotyping batch, the first two principal components, white blood cell count and platelet count, body mass index, physical activity, smoking status, alcohol consumption frequency, blood pressure and blood pressure-lowering medication, cholesterol, triglycerides and lipid-lowering medication, sleep duration and insomnia, type 2 diabetes status, and familial history of cardiovascular disease) and corresponding 95% confidence intervals (gray shadowed area) using restricted cubic splines for mtDNA copy number with knots at distribution of 5th, 50th, and 95th percentiles. The density on the right y-axis shows distribution of baseline mtDNA copy number. Since mtDNA copy number was scaled before analyses, the distribution is normal. CAD: coronary artery disease; HF: heart failure.

**Figure 4.**
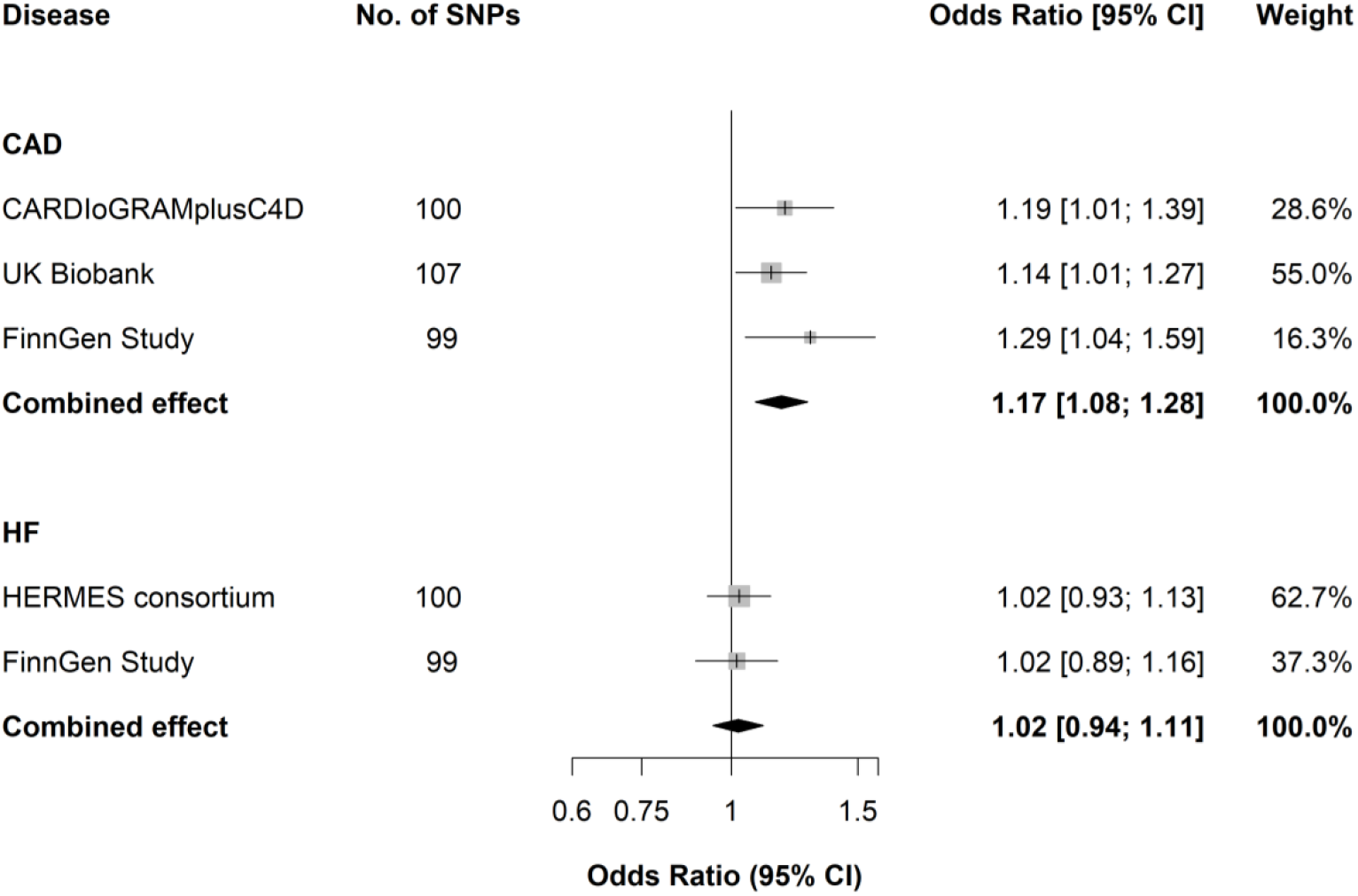
Mendelian Randomization study of mtDNA copy number on the risk of CAD and HF (Instruments retrieved from Longchamps *et al*.) Estimated ORs for the effect of per-SD decrease in mtDNA copy number on CAD and HF, obtained from an MR inverse-variance weighted method, per outcome database separately and combined over the different databases using fixed-effect meta-analyses. CARDIoGRAMplusC4D: Coronary Artery Disease Genome-Wide Replication and Meta-analysis plus the Coronary Artery Disease Genetics; HERMES consortium: Heart Failure Molecular Epidemiology for Therapeutic Targets consortium. UK Biobank data of heart failure was already integrated into HERMES consortium.

#### Sensitivity analysis

We observed no evidence favoring an interaction between mtDNA-CN and sex (p for interaction = 0.2 for CAD, 0.7 for HF); in line, in sex-stratified analyses, the estimates between men and women were similar (**Table S3**). Interaction was observed between mtDNA-CN and age at baseline for CAD (p for interaction < 0.001). After stratification by age groups, HRs obtained from model 2 for CAD slightly attenuated from younger group (<50 years) to older group (50∼60 years and >60 years) (HR: 1.06, 1.04 and 1.02, respectively) (**Table S4**). However, no interaction was detected between mtDNA-CN and age at baseline for HF (p for interaction = 0.2); though HR in the younger group was also higher for HF, this may be due to the very limited number of cases in this group

When analyses were conducted for MI and IHD separately, cumulative incidences were higher in lower quintiles compared with the highest quintile (**Figure S2**) for MI and IHD; estimates from cox proportional hazard regression models did not differ considerably from when all CADs were considered (**Table S2-S4**).

In addition, missing data in covariates were present (**Table S5**), and 162,002 (59%) of 273,619 individuals included in the current study provided complete data for all variables. The absolute difference in the baseline characteristics between these participants with and without complete data was very limited (**Table S6**). Furthermore, the main results from sensitivity analyses restricting to complete cases did not differ materially from the results obtained after imputation (**Table S7**).

### Mendelian Randomization

#### Main analyses

For the included 129 genetic instrumental variables, 4 of which with an F-statistics below 10 were discarded to avoid weak instrumental bias. F-statistics for each of the remaining 125 SNPs were higher than 10 and ranged from 15.6 to 634.4, and a total of 2.0% variation were explained by the instruments (107 non-palindromic SNPs at most included in the MR analyses) (**Table S8**).

For CAD, the pooled OR of the primary IVW estimates from CARDIoGRAMplusC4D, UKB, and FinnGen of a one-SD decrease in mtDNA-CN was 1.17 (95%CI: 1.08, 1.28). Estimates from WME and MR-Egger generally did not differ substantially with the exception of UKB where the point estimates attenuated to some extent. No pleiotropy was detected by the intercept of MR-Egger (p > 0.05). Though outliers were identified by MR PRESSO in each database, estimates after outlier removal remained similar to those obtained from IVW (**Table S9**).

For HF, the combined OR from IVW obtained in the HERMES consortium and FinnGen of per one-SD decrease in mtDNA-CN was 1.02 (95%CI: 0.94, 1.11). Results from WME were similar, and we observed no evidence for horizontal pleiotropy from MR-Egger intercept (p > 0.05); outliers were spotted in the HERMES consortium assessed by MR PRESSO, but outlier-corrected estimates did not materially differ with those generated from IVW (**Table S10**).

#### Sensitivity analyses

When we used genetic instrumental variables from Hägg *et al*., F statistics for each of the 66 SNPs were higher than 10 and ranged from 29.9 to 277.4, and a total of 1.3% variation were explained by the instruments (58 non-palindromic SNPs at most included in the MR analyses). Detailed full information of the instruments was in **Table S11**. A one-SD decrease in mtDNA-CN was associated with 1.16-fold (95%CI:1.05, 1.27), 1.00-fold (95%CI:0.90, 1.10) higher risk of CAD and HF in meta-analysis, respectively (**Figure S3**). MR sensitivity analyses including WME, MR-Egger, and MR PRESSO were presented in **Table S12-13**.

## 4. Discussion

In the present study, we used both a prospective cohort study design and MR study design to assess the relationship of mtDNA-CN with the risk of incident CAD and HF. Results from the multivariable adjusted prospective analyses suggested an association between lower mtDNA-CN with higher risk of CAD and HF, whereas findings from MR analyses only confirmed an association between genetically predicted lower mtDNA-CN with a higher risk of CAD, but not on the risk of HF, possibly reflecting evidence of causality for CAD.

Consistent with the results found in our study, previous studies all showed that lower mtDNA-CN measured from peripheral blood was related to an increased risk of CVD (4-11). The only prospective study that assessed the relationship between mtDNA content and either CAD or HF used the Atherosclerosis Risk in Communities (ARIC) Study (9,10). In the ARIC study, composed of 20,163 participants (2460 incident CHD) with a mean follow-up of 13.5 years, a lower mtDNA-CN was associated with an increased risk of incident CHD. Similarly, with 10,802 participants (2227 incident HF cases) followed-up for a mean of 23.1 years, lower mtDNA-CN was linked to an increased risk of HF. Residual confounding, in particular factors relevant to both mitochondrial function and CVD such as physical activity and sleep, was not taken into account. However, in our multivariable-adjusted analysis, additional adjustment for these covariates did not further attenuate the estimates substantially.

To the best of our knowledge, the current study is the first to evaluate whether the association between mtDNA-CN and risk of CVD is causal using an MR approach. MR analyses with the genetic instruments for mtDNA-CN confirmed the detrimental effect of lower mtDNA content on the risk of CAD observed in the cohort study. The effect size from the MR study (OR: 1.16 [95%CI: 1.05, 1.27]) is larger than that in the cohort study (HR for continuous analyses: 1.03 [95%CI: 1.01, 1.04]), though comparisons should be elaborated with caution as the effect from genetic predisposition is assumed throughout the whole lifespan while the effect in the cohort sustains merely during the follow-up. However, evidence in our study suggests that a lifelong expose to a low mtDNA-CN would lead to an elevated risk of CAD. Nevertheless, several factors merit thoughtful consideration in terms of the interpretation of the null effect on HF in MR analyses. HF has substantial phenotypic heterogeneity, which can be defined by ejection fraction (EF) and diastolic function; more than half of patients have preserved EF while over 40% cases have isolated diastolic dysfunction (29). Moreover, a large degree of variation has been described even within patients with preserved EF (30,31). However, stratification by cause of HF in the UKB ended up with a low number of cases and insufficient statistical power. In addition, cause-specific GWAS summary-level data of HF are currently not available. Considering that the estimates in the MR studies are completely null on HF in either single database or meta-analysis, it is likely that the effect of mtDNA-CN on different subtypes of HF, if this exists at all, would be very small.

### Study strengths and limitations

The main strength of our study is that we adopted the triangulation of causal inference in etiological epidemiology described by Lawlor *et al*. (12). Two different approaches were simultaneously used to determine the effect of mitochondrial content on CVD, which include multivariable regression analyses in a prospective cohort study and refinements of exposure by MR design. The consistency between biochemically measured and genetically determined mtDNA-CN in relation to CAD increased the credibility of the results. Given that the absence of randomized clinical trials with respect to mtDNA-CN and CAD so far, the analyses that have been performed in the present study provide the foremost evidence on the association between mtDNA content and CAD. Other important strengths of our prospective cohort study include the large sample size as well as the considerable number of incident cases from UKB, comprehensive assessment of confounding factors, and subtype analyses of MI and IHD within CAD. In MR studies, we meta-analyzed three large databases from which SNP-outcome associations were derived, comprising a substantial size of overall participants and cases. The results are quite consistent across different databases, and the precision of the pooled MR estimates obtained from different databases increased significantly.

Several limitations should also be acknowledged. First, mtDNA content was assessed in peripheral blood cells, which may be different from cells in vasculature or in the heart. Nevertheless, it has been shown that within individuals, the blood cell and cardiomyocyte mtDNA-CN were significantly correlated, with a Pearson correlation coefficient of 0.72 (32). In addition, the initial calculation of mtDNA-CN from chip arrays might have introduced noise due to the small number of variants. To this end, a weighted mtDNA-CN was implemented, which approximates what would be estimated from exome sequencing and has been validated previously (16). Second, despite over 100 instrumental variables by Longchamps *et al* were used from in the main MR analyses, the variation of mtDNA-CN explained by theses SNPs was very small. In the study, authors claimed less than 1% of the variance in mtDNA-CN explained by GWAS loci when predicted into the ARIC cohort. Therefore, they were unable to determine the directionality of the effect between mtDNA-CN and outcomes since the MR studies might be unpowered. Moreover, the genetic variants associated with mtDNA-CN are likely to be highly pleiotropic (19). Notwithstanding, using the online power calculation for MR (https://github.com/kn3in/mRnd) (33), we had a power of 0.8 to detect an OR of 1.10 to 1.35 with an assumed explained variation of 0.5% (**Figure S4A**), which is much smaller than the estimated variance of 1% by Longchamps *et al*., despite the calculated total variation was 2.0% as described in the methods section. Similarly, we had 80% power to detect an OR of 1.1 to 1.2 in all enrolled databases with an explained variation of 1.3% by the genetic instruments in the sensitivity analyses of the MR study (**Figure S4B**). While we acknowledge the possibility of pleiotropic effects of included genetic instruments, this is likely to be vertical. There is no horizontal pleiotropic effect detected in the MR-Egger intercept analyses; although horizontal pleiotropy was detected via MR PRESSO global test, but only restricted to the analyses with outliers (**Table S9-S10**). Additionally, in spite of the significant associations with a substantial number of quantitative traits in the study from Longchamps *et al*., the associations of mtDNA-CN with conventional CVD risk factors, including but not limited to obesity, unfavorable lipid profiles, and blood pressure are in the same direction as the associations identified in our study with both CAD and HF, which further strengthens the notion that these might be the mediators in the causal pathway from mtDNA content to CVD. Third, since the population of non-European are highly heterogenous in UKB, we restricted the prospective analyses to White European populations; furthermore, MR analyses were also performed predominantly in European-descent individuals, apart from about 23% individuals with a non-European background in CARDIoGRAMplusC4D. It is therefore maybe inappropriate to extrapolate our findings to other populations with different ethnic backgrounds. Further studies in non-European subjects are needed to confirm the effect of mtDNA on CVD.

## 5. Conclusion

This study provides the first evidence of a possible causal association between mtDNA-CN and the risk of CAD. Further studies are required to fully understand how mtDNA affect atherogenic risk development.

## Data Availability

The data we used are either from publically available sources as indicated in the manuscript, or from the UKB, which is open to all researches at reasonable request.

## Acknowledgements

The present study has been conducted using the UK Biobank Resource (Application Number 56340) that is available to researchers. The authors acknowledge the participants and investigators of all consortia that contributed summary statistics data, including the UK Biobank, CARDIoGRAMplusC4D, HERMES consortium, and the FinnGen study.

