## Supplementary materials for "Low mitochondrial copy number drives atherogenic cardiovascular disease: evidence from prospective cohort analyses in the UK Biobank combined with Mendelian Randomization"

### Supplementray files

**Supplement texts**

**Table S1** GWAS data sources for Mendelian Randomization

**Table S2** Hazard ratios of incident CAD and HF by quintiles of mtDNA copy number

**Table S3** Hazard ratios of incident CAD and HF by levels of mtDNA copy number, stratified by sex

**Table S4** Hazard ratios of incident CAD and HF by levels of mtDNA copy number, stratified by age

**Table S5** Number and frequencies of missingness in each variable (N = 273,619)

**Table S6** Differences between participants with complete data and participants with missing data

**Table S7** Cox proportional hazard regression in complete cases

**Table S8** Genetic instruments at genome-wide significant level for mtDNA copy number in the main MR analyses (f Instruments retrieved rom Longchamps et al.)

**Table S9** Mendelian Randomization results of mtDNA copy number on the risk of CAD (Instruments retrieved from Longchamps et al.)

**Table S10** Mendelian Randomization results mtDNA copy number on the risk of HF (Instruments retrieved retrieved from Longchamps et al.)

**Table S11** Genetic instruments at genome-wide significant level for mtDNA copy number in the sensitivity MR analysis (Instruments retrieved from Hägg et al.)

**Table S12** Mendelian Randomization results of mtDNA copy numberon on the risk of CAD (Instruments retrieved retrieved from Hägg et al.)

**Table S13** Mendelian Randomization results mtDNA copy numberon on the risk of HF (Instruments retrieved from Hägg et al.)

**Figure S1** Flowchart of participants inclusion in UK Biobank

**Figure S2** Cumulative Incidence for competing risk of MI and IHD by quintiles of mtDNA mtDNA copy number

**Figure S3** Mendelian Randomization study of mtDNA copy number on the risk of CAD and HF (Instruments retrieved from Hägg et al.)

**Figure S4** Statistical power of Mendelian Randomization analyses

### Supplement texts

**Mendelian Randomization**

*Outcome data source*

CARDIoGRAMplusC4D (Coronary Artery Disease Genome-Wide Replication and Meta-analysis plus the Coronary Artery Disease Genetics) represents a collaborative effort to combine data from multiple large scale genetic studies to identify risk loci for CAD (coronary artery disease), which assembled 60,801 cases and 123,504 control subjects for 48 studies, of which 77% of the participants were of European ancestry, 19% were of south and east Asian ancestry, and a small proportion were Hispanic and African Americans. CAD cases were identified as an inclusive diagnosis of myocardial infarction, acute coronary syndrome, chronic stable angina, or coronary stenosis >50% (1,2).

HERMES Consortium (Heart Failure Molecular Epidemiology for Therapeutic Targets Consortium) is an international collaboration to investigate the genetic basis of HF (heart failure), which comprises 51 population-based cohorts, case-control studies and randomized clinical trials, including over 65,000 heart failure cases. Summary statistics used in the current study includes 26 studies (including data from the UK Biobank) with 47,309 heart failure cases and 930,014 controls of European ancestry (3). HF assessment was performed by at least one of the either discharge registries, cause of death registries or physician adjudication/ diagnosis in each cohort.

The UK Biobank cohort has been described in the previous text. We restricted the analyses to the participants of European ancestry, and who were in the full released imputed genomics databases. Disease diagnoses were coded according to the International Classification of Diseases (ICD) and were retrieved from linkage with NHS database. CAD cases were defined as angina pectoris (I20), myocardial infarction (I21 and I22), and acute and chronic ischemic heart disease (I24 and I25). In total, 52,946 cases and 446,495 control subjects were identified. We performed linear mixed regression analyses to assess the associations between genetic instruments and CAD, adjusted for age, sex and 10 principal components, genotype batch and corrected for familial relationship using BOLD_LMM (v2.3.2).

The FinnGen study is an ongoing cohort study launched in 2017, which included the genetic data generated from biobank samples and health-related data from social and healthcare registers. In the FinnGenn study, GWAS were performed across a broad spectrum of phenotypes, adjusted for age, sex, principal components and genotype batch effect. Detailed information regarding participants for GWAS, genotype platforms and statistical analysis protocols are available at FinnGenn website (<https://www.finngen.fi/en/>). International Statistical Classification of Diseases and Related Health Problems (ICD) coded hospital discharge or death are used to define cases in the FinnGenn study. Major coronary heart disease was defined as angina pectoris (I20), myocardial infarction (I21 to I23), ischemic heart diseases (I24 and I25), cardiac arrest (I46), and other unattended or cause unknown sudden death (R96 and R98).

### Table S1 GWAS data sources for Mendelian Randomization

| **Consortium** | **Phenotype** | **Cases/**  **Controls** | **Population** | **Phenotype definition** | **Covariates** |
| --- | --- | --- | --- | --- | --- |
| CARDIoGRAMplus  C4D  (Nikpay et al. 2015, Nat Genet) | CAD | 60,801  123,504 | European 77%, South Asian 13%, East Asian 6% and a small proportion Hispanic and African Americans (48 cohorts) | Inclusive diagnosis of myocardial infarction, acute coronary syndrome, chronic stable angina, or coronary stenosis > 50% | Analyses were performed within each included cohort |
| UK Biobank^*^ | CAD | 52,946 446,495 | British (European ancestry) | ICD-10 (I20 to I22, I24 and I25) | Age, sex and 10 PCs and genotyping batch |
| HERMES  (Shah et al. 2020, Nat Commun) | HF | 47,309  930,014 | European (26 cohorts with 29 datasets, including data from UKB). | A clinical diagnosis of HF of any etiology with no inclusion criteria based on LV ejection fraction | Age, sex and PCs within each cohort |
| FinnGen Study  (Data freeze 5, 2021 May 11) | CHD | 21,012  197,780 | Finnish | Hospital discharge or cause of death of ICD-10 (I20 to I25, I46, R96 and R98) and ICD-9/ICD-8 (410 and 4110) | Age, sex, 10 PCs and genotyping batch |
|  | HF | 23,397  194,811 |  | Hospital discharge or cause of death of ICD-10 (I11.0, I13.0, I13.2, I50) or ICD-9 (4029B\|428) or ICD-8 (42700\|42710\|428\|7824), or KELA codes (201) or medicine purchases (C03CA01\|C03EB01) |  |

CARDIoGRAMplusC4D: Coronary Artery Disease Genome-Wide Replication and Meta-analysis plus the Coronary Artery Disease Genetics; HERMES consortium: Heart Failure Molecular Epidemiology for Therapeutic Targets consortium; CAD: coronary artery disease; HF: heart failure; CHD: coronary heart disease; PCs: Principal components; ICD: International Statistical Classification of Diseases and Related Health Problems.

^*^UK Biobank data of heart failure was already integrated into HERMES consortium.

### Table S2 Hazard ratio of incident CAD and HF by quintiles of mtDNA copy number

|  | **Continuous**  **(N = 273,619)** | **Q1 (N = 54,724)** | **Q2 (N = 54,724)** | **Q3 (N = 54,724)** | **Q4 (N = 54,724)** | **Q5 (N = 54,724)** |
| --- | --- | --- | --- | --- | --- | --- |
| **CAD** |  |  |  |  |  |  |
| Incident cases | 18,346 (6.7%) | 4125 (7.5%) | 3959 (7.2%) | 3618 (6.6%) | 3504 (6.4%) | 3140 (5.7%) |
| Model 1 | 1.06 (1.05, 1.08) | 1.18 (1.13, 1.24) | 1.17 (1.12, 1.23) | 1.10 (1.05, 1.15) | 1.09 (1.04, 1.14) | 1.0 (reference) |
| Model 2 | 1.03 (1.01, 1.04) | 1.08 (1.03, 1.14) | 1.09 (1.04, 1.15) | 1.04 (0.99, 1.09) | 1.06 (1.01, 1.11) | 1.0 (reference) |
| **HF** |  |  |  |  |  |  |
| Incident cases | 5795 (2.1%) | 1395 (2.5%) | 1235 (2.3%) | 1151 (2.1%) | 1062 (1.9%) | 952 (1.7%)) |
| Model 1 | 1.09 (1.06, 1.12) | 1.28 (1.17, 1.39) | 1.18 (1.08, 1.28) | 1.13 (1.04, 1.24) | 1.09 (1.00, 1.19) | 1.0 (reference) |
| Model 2 | 1.05 (1.02, 1.08) | 1.15 (1.05, 1.24) | 1.08 (0.99, 1.18) | 1.07 (0.98, 1.16) | 1.05 (0.96, 1.14) | 1.0 (reference) |
| **MI** |  |  |  |  |  |  |
| Incident cases | 5535 (2.0%) | 1229 (2.2%) | 1186 (2.2%) | 1139 (2.1%) | 1035 (1.9%) | 946 (1.7%) |
| Model 1 | 1.07 (1.04, 1.09) | 1.19 (1.09, 1.29) | 1.18 (1.08, 1.28) | 1.16 (1.06, 1.26) | 1.07 (0.98, 1.17) | 1.0 (reference) |
| Model 2 | 1.02 (0.99, 1.05) | 1.06 (0.97, 1.15) | 1.08 (0.99, 1.17) | 1.08 (0.99, 1.18) | 1.03 (0.94, 1.12) | 1.0 (reference) |
| **IHD** |  |  |  |  |  |  |
| Incident cases | 15,441 (5.6%) | 3490 (6.4%) | 3360 (6.1%) | 3026 (5.5%) | 2952 (5.4%) | 2613 (4.8%) |
| Model 1 | 1.06 (1.05, 1.08) | 1.19 (1.13, 1.25) | 1.19 (1.13, 1.25) | 1.10 (1.04, 1.16) | 1.10 (1.05, 1.16) | 1.0 (reference) |
| Model 2 | 1.03 (1.01, 1.05) | 1.09 (1.03, 1.14) | 1.10 (1.05, 1.16) | 1.04 (0.99, 1.10) | 1.06 (1.01, 1.12) | 1.0 (reference) |

Estimated hazard ratios for the effect of per-SD decrease in mtDNA copy number (continuous), or for the 1st to the 4th quintile compared to the 5th (reference) quintile (categorical) on CAD, HF, MI or IHD.

Model 1 was adjusted for age, sex, genotyping batch, the first two principal components, white blood cell count and platelet count.

Model 2 was model 1 additionally adjusted for body mass index, physical activity, smoking status, alcohol consumption frequency, blood pressure and blood pressure-lowering medication, cholesterol, triglycerides and lipid-lowering medication, sleep duration and insomnia, type 2 diabetes status, and familial history of cardiovascular disease.

CAD: coronary artery disease; HF: heart failure; MI: myocardial infarction; IHD: ischemic heart disease.

### Table S3 Hazard ratio of incident CAD and HF by levels of mtDNA copy number, stratified by sex

|  | **CAD** | **HF** | **MI** | **IHD** |
| --- | --- | --- | --- | --- |
| **Female (N = 150,888)** | | | | |
| Incident cases | 6733 (4.5%) | 2295 (1.5%) | 1707 (1.1%) | 5202 (3.4%) |
| Model 1 | 1.04 (1.02, 1.07) | 1.09 (1.05, 1.14) | 1.05 (1.00, 1.10) | 1.05 (1.02, 1.07) |
| Model 2 | 1.01 (0.98, 1.03) | 1.04 (1.00, 1.09) | 1.00 (0.95, 1.05) | 1.01 (0.98, 1.03) |
| **Male (N = 122,731)** | | | | |
| Incident cases | 11,613 (9.5%) | 3500 (2.9%) | 3828 (3.1%) | 10,239 (8.3%) |
| Model 1 | 1.07 (1.05, 1.09) | 1.08 (1.05, 1.12) | 1.07 (1.04, 1.11) | 1.07 (1.05, 1.09) |
| Model 2 | 1.04 (1.02, 1.06) | 1.05 (1.01, 1.09) | 1.03 (1.00, 1.06) | 1.04 (1.02, 1.06) |
| **P for interaction^*^** | 0.2 | 0.7 | 0.7 | 0.3 |

Hazard ratios are per one-SD decrease in mtDNA copy number.

Model 1 was adjusted for age, sex, genotyping batch, the first two principal components, white blood cell count and platelet count.

Model 2 was model 1 additionally adjusted for body mass index, physical activity, smoking status, alcohol consumption frequency, blood pressure and blood pressure-lowering medication, cholesterol, triglycerides and lipid-lowering medication, sleep duration and insomnia, type 2 diabetes status, and familial history of cardiovascular disease.

The incident cases of MI and IHD does not add up to the cases of CAD since some participants contribute data to both MI and IHD, for example, the diagnosis of MI and IHD occurred on the same day.

^*^ P value for interaction was derived from model 2 adding extra multiplicative terms of sex and mtDNA copy number.

CAD: coronary artery disease; HF: heart attack; MI: myocardial infarction; IHD: ischemic heart disease.

### Table S4 Hazard ratio of incident CAD and HF by levels of mtDNA copy number, stratified by age

|  | **CAD** | **HF** | **MI** | **IHD** |
| --- | --- | --- | --- | --- |
| **< 50 years (N = 63,277)** | | | | |
| Incident cases | 1666 (2.6%) | 308 (0.5%) | 600 (0.9%) | 1334 (2.1%) |
| Model 1 | 1.14 (1.08, 1.19) | 1.24 (1.01, 1.39) | 1.10 (1.02, 1.2) | 1.15 (1.09, 1.22) |
| Model 2 | 1.06 (1.01, 1.12) | 1.15 (1.02, 1.29) | 1.02 (0.94, 1.11) | 1.07 (1.02, 1.14) |
| **50~60 years (N = 104,209)** | | | | |
| Incident cases | 10,627 (10.2%) | 3966 (3.8%) | 3001 (2.9%) | 9082 (8.7%) |
| Model 1 | 1.09 (1.06, 1.12) | 1.08 (1.02, 1.13) | 1.09 (1.04, 1.14) | 1.10 (1.06, 1.13) |
| Model 2 | 1.04 (1.01, 1.06) | 1.02 (0.97, 1.07) | 1.03 (0.98, 1.08) | 1.04 (1.01, 1.07) |
| **> 60 years (N = 106,133)** | | | | |
| Incident cases | 6053 (5.7%) | 1521 (1.4%) | 1934 (1.8%) | 5025 (4.7%) |
| Model 1 | 1.05 (1.03, 1.07) | 1.10 (1.07, 1.13) | 1.05 (1.02, 1.09) | 1.05 (1.03, 1.07) |
| Model 2 | 1.02 (1.00, 1.04) | 1.06 (1.03, 1.10) | 1.02 (0.98, 1.06) | 1.02 (1.00, 1.04) |
| **P for interaction^*^** | < 0.001 | 0.2 | 0.7 | <0.001 |

Hazard ratios are per one-SD decrease in mtDNA copy number.

Model 1 was adjusted for age, sex, genotyping batch, the first two principal components, white blood cell count and platelet count.

Model 2 was model 1 additionally adjusted for body mass index, physical activity, smoking status, alcohol consumption frequency, blood pressure and blood pressure-lowering medication, cholesterol, triglycerides and lipid-lowering medication, sleep duration and insomnia, type 2 diabetes status, and familial history of cardiovascular disease.

The incident cases of MI and IHD does not add up to the cases of CAD since some participants contribute data to both MI and IHD, for example, the diagnosis of MI and IHD occurred on the same day.

^*^ P value for interaction was derived from model 2 adding extra multiplicative terms of age and mtDNA copy number.

CAD: coronary artery disease; HF: heart attack; MI: myocardial infarction; IHD: ischemic heart disease.

### Table S5 Number and frequencies of missingness in each variable (N = 273,619)

| **Variable** | **Number (Percentage)** |
| --- | --- |
| White blood cell count | 8149 (3%) |
| Platelet count | 8145 (3%) |
| BMI (kg/m^2^) | 782 (0.3%) |
| Deprivation index | 331 (0.1%) |
| Blood pressure | 15,594 (5.7%) |
| Cholesterol | 12,689 (4.6%) |
| LDL | 13,168 (4.8%) |
| Triglycerides | 12,903 (4.7%) |
| Physical activity (MET of moderate-vigorous per week) | 51,163 (18.7%) |
| Alcohol consumption frequency | 177 (0.1%) |
| Smoking status | 894 (0.3%) |
| Insomnia | 180 (0.1%) |
| Familial history of CVD | 25,564 (9.3%) |

Covariates do not have any missingness were not presented in the table, including age, sex, principal components, genotyping batch, sleep duration, blood-lowering medication and lipid-lowering medication, and type 2 diabetes mellitus; BMI: Body mass index; LDL: low-density lipoprotein cholesterol; CVD: cardiovascular disease.

### Table S6 Differences between participants with complete data and participants with missing data

| **Variable** | **Complete cases**  (N = 162,002) | **Participants with missingness**  (N = 111,617) |
| --- | --- | --- |
| mtDNA copy number | 0 (1.0) | 0 (1.0) |
| CAD incident cases | 10,495 (6.5%) | 7851 (7.0%) |
| CAD follow up time (years) | 11.8 (10.9, 2.4) | 11.9 (11.1, 12.6) |
| HF incident cases | 3147 (1.9%) | 2468 (2.4%) |
| HF follow up time (years) | 11.8 (11.1, 12.5) | 11.9 (11.2, 12.6) |
| White blood cell count | 6.8 (2.0) | 6.9 (2.0) |
| Platelet count | 252.5 (58.9) | 256.0 (60.3) |
| Age (years) | 56.2 (8.0) | 56.9 (8.0) |
| BMI (kg/m^2^) | 27.1 (4.6) | 27.5 (4.9) |
| Deprivation index | -1.8 (2.8) | -1.4 (3.0) |
| Diastolic blood pressure (mmHg) | 82.3 (10.1) | 82.5 (10.1) |
| Systolic blood pressure (mmHg) | 137.7 (18.5) | 138.9 (18.9) |
| Blood pressure-lowering medication (yes, %) | 27,255 (16.8 %) | 20,867 (18.7%) |
| Total cholesterol (mmol/L) | 5.8 (1.1) | 5.8 (1.1) |
| HDL (mmol/L) | 1.5 (0.4) | 1.5 (0.4) |
| LDL (mmol/L) | 3.6 (0.8) | 3.6 (0.9) |
| Triglycerides (mmol/L) | 1.7 (1.0) | 1.8 (1.0) |
| Cholesterol lowering medication | 20,585 (12.7%) | 15,038 (13.5%) |
| Physical activity (moderate-vigorous MET hours/week) | 26.9 (33.8) | 27.2 (34.7) |
| Alcohol consumption frequency (Twice or less per week) | 83760 (51.7%) | 63525 (57.0%) |
| Smoking status (ex-smoker) | 55,743 (34.4%) | 37,610 (34.0%) |
| Sleep duration (hours) | 7.2 (1.1) | 7.1 (1.4) |
| Insomnia (never/rarely) | 41,219 (25.4%) | 25,165 (22.6%) |
| Familial history of CVD (yes) | 69,601 (43.0%) | 38,390 (44.6%) |

Data are mean (SD) or median (interquartile range, IQR) for continuous variables and frequency (percentage) for categorical variables. All differences are statistically significant (p<0.05).

CAD: coronary artery disease; BMI: Body mass index; LDL: low-density lipoprotein cholesterol; CVD: cardiovascular disease.

### Table S7 Cox proportional hazard regression in complete cases

|  | **Continuous^*^**  **(N = 162,002)** | **Q1 (N = 32,401)** | **Q2 (N = 32,401)** | **Q3 (N = 32,400)** | **Q4 (N = 32,400)** | **Q5 (N = 32,400)** |
| --- | --- | --- | --- | --- | --- | --- |
| **CAD** |  |  |  |  |  |  |
| Incident cases | 10,495 (6.5%) | 2356 (7.3%) | 2287 (7.1%) | 2081 (6.4%) | 1989 (6.1%) | 1782 (5.5%) |
| Model 1 | 1.06 (1.04, 1.08) | 1.19 (1.12, 1.27) | 1.20 (1.13, 1.28) | 1.11 (1.04, 1.18) | 1.09 (1.02, 1.16) | 1.0 (Reference) |
| Model 2 | 1.03 (1.01, 1.05) | 1.10 (1.04, 1.17) | 1.13 (1.06, 1.20) | 1.06 (1.00, 1.13) | 1.05 (0.99, 1.12) | 1.0 (Reference) |
| **HF** |  |  |  |  |  |  |
| Incident cases | 3147 (1.9%) | 754 (2.3%) | 690 (2.1%) | 618 (1.9%) | 560 (1.7%) | 525 (1.6%) |
| Model 1 | 1.09 (1.05, 1.13) | 1.24 (1.11, 1.39) | 1.19 (1.07, 1.34) | 1.10 (0.98, 1.24) | 1.03 (0.92, 1.17) | 1.00 (Reference) |
| Model 2 | 1.05 (1.01, 1.09) | 1.12 (1.00, 1.26) | 1.10 (0.98, 1.23) | 1.04 (0.92, 1.17) | 1.00 (0.89, 1.12) | 1.00 (Reference) |
| **MI** |  |  |  |  |  |  |
| Incident cases | 8851 (5.5%) | 669 (2.1%) | 656 (2.0%) | 654 (2.0%) | 606 (1.9%) | 530 (1.6%) |
| Model 1 | 1.05 (1.01, 1.09) | 1.19 (1.12, 1.27) | 1.2 (1.13, 1.28) | 1.11 (1.04, 1.18) | 1.09 (1.02, 1.16) | 1.0 (Reference) |
| Model 2 | 1.01 (0.98, 1.05) | 1.05 (0.94, 1.18) | 1.08 (0.96, 1.21) | 1.12 (1.00, 1.25) | 1.07 (0.96, 1.21) | 1.0 (Reference) |
| **IHD** |  |  |  |  |  |  |
| Incident cases | 3115 (1.9%) | 1994 (6.2%) | 1955 (6.0%) | 1737 (5.4%) | 1672 (5.2%) | 1493 (4.6%) |
| Model 1 | 1.06 (1.04, 1.09) | 1.19 (1.11, 1.27) | 1.22 (1.14, 1.30) | 1.10 (1.03, 1.18) | 1.09 (1.02, 1.17) | 1.0 (Reference) |
| Model 2 | 1.03 (1.01, 1.06) | 1.10 (1.03, 1.17) | 1.14 (1.06, 1.22) | 1.05 (0.98, 1.13) | 1.05 (0.98, 1.13) | 1.0 (Reference) |

Model 1 was adjusted for age, sex, genotyping batch, the first two principal components, white blood cell count and platelet count.

Model 2 was model 1 additionally adjusted for body mass index, physical activity, smoking status, alcohol consumption frequency, blood pressure and blood pressure-lowering medication, cholesterol, triglycerides and lipid-lowering medication, sleep duration and insomnia, type 2 diabetes status, and familial history of cardiovascular disease.

^*^Continuous hazard ratio is per one-SD decrease in mtDNA copy number.

CAD: coronary artery disease; HF: heart failure; MI: myocardial infarction; IHD: ischemic heart disease.

### Table S8 Genetic instruments at genome-wide significant level for mtDNA copy number in the main MR analyses (Instruments retrieved from Longchamps *et al.*)

| **SNP** | **EA/Non-EA** | **EAF** | **Beta** | **se** | **F statistics** | **PVE (%)** |
| --- | --- | --- | --- | --- | --- | --- |
| rs1569419 | T/C | 0.23 | -0.02 | 0.003 | 84.0 | 0.019 |
| rs3818157 | G/A | 0.54 | 0.02 | 0.002 | 99.8 | 0.022 |
| rs204071 | C/T | 0.93 | -0.02 | 0.004 | 29.3 | 0.006 |
| 1:156455314 | CTT/C | 0.32 | 0.01 | 0.002 | 41.2 | 0.009 |
| rs6425521 | C/A | 0.20 | -0.02 | 0.003 | 67.7 | 0.015 |
| rs9425311 | G/T | 0.51 | 0.01 | 0.002 | 40.4 | 0.009 |
| 1:205246482 | TTTTG/T | 0.61 | 0.02 | 0.002 | 57.4 | 0.013 |
| rs10749636 | G/A | 0.24 | -0.02 | 0.002 | 38.3 | 0.009 |
| rs655029 | G/A | 0.29 | 0.01 | 0.002 | 36.2 | 0.008 |
| rs711244 | C/T | 0.58 | 0.01 | 0.002 | 42.1 | 0.009 |
| rs2302643 | G/A | 0.55 | -0.01 | 0.002 | 32.5 | 0.007 |
| rs151084028 | T/G | 0.99 | 0.09 | 0.014 | 34.8 | 0.009 |
| rs865551 | C/G | 0.36 | 0.02 | 0.002 | 52.5 | 0.012 |
| rs62641680 | G/A | 0.97 | 0.11 | 0.006 | 293.9 | 0.064 |
| rs74874677 | A/G | 0.98 | 0.10 | 0.007 | 201.9 | 0.044 |
| rs12052715 | C/G | 0.28 | 0.02 | 0.002 | 51.1 | 0.011 |
| rs147820690 | C/T | 1.00 | -0.12 | 0.021 | 36.3 | 0.008 |
| rs78909033 | G/A | 0.87 | 0.02 | 0.003 | 64.0 | 0.014 |
| rs13084580 | C/T | 0.89 | -0.03 | 0.003 | 66.1 | 0.015 |
| rs6786055 | G/T | 0.37 | 0.01 | 0.002 | 45.6 | 0.010 |
| rs1354034 | T/C | 0.40 | 0.03 | 0.002 | 211.9 | 0.046 |
| rs6778131 | T/A | 0.38 | 0.01 | 0.002 | 30.5 | 0.007 |
| rs1420476 | T/A | 0.04 | -0.02 | 0.005 | 20.6 | 0.005 |
| rs34894010 | C/G | 0.96 | -0.04 | 0.005 | 51.7 | 0.011 |
| rs755492124 | TAAAG/T | 0.61 | -0.01 | 0.002 | 41.0 | 0.009 |
| rs6894574 | T/C | 0.33 | 0.01 | 0.002 | 25.8 | 0.006 |
| rs2736100 | C/A | 0.50 | 0.02 | 0.002 | 67.7 | 0.015 |
| rs34592828 | G/A | 0.96 | -0.03 | 0.005 | 40.4 | 0.009 |
| rs114694170 | T/C | 0.94 | -0.04 | 0.005 | 65.6 | 0.015 |
| rs56116444 | T/G | 0.92 | 0.03 | 0.004 | 39.5 | 0.009 |
| rs193541 | C/T | 0.42 | -0.01 | 0.002 | 37.7 | 0.008 |
| rs926326 | A/G | 0.23 | -0.02 | 0.003 | 46.7 | 0.010 |
| rs2844484 | A/G | 0.39 | 0.02 | 0.002 | 65.2 | 0.014 |
| rs45552734 | C/T | 0.88 | -0.02 | 0.003 | 41.6 | 0.009 |
| rs511515 | A/G | 0.30 | -0.02 | 0.002 | 83.4 | 0.018 |
| rs5745582 | C/T | 0.82 | -0.03 | 0.003 | 87.4 | 0.019 |
| rs4895441 | A/G | 0.73 | -0.02 | 0.002 | 71.8 | 0.016 |
| rs7744765 | T/C | 0.59 | 0.02 | 0.002 | 51.1 | 0.011 |
| rs200957609 | G/A | 1.00 | -0.12 | 0.021 | 29.7 | 0.007 |
| rs35585318 | T/C | 0.61 | 0.01 | 0.002 | 27.7 | 0.006 |
| rs6943701 | A/T | 0.82 | -0.02 | 0.003 | 43.2 | 0.010 |
| rs17260734 | T/A | 0.52 | -0.01 | 0.002 | 28.3 | 0.006 |
| rs445 | C/T | 0.90 | -0.02 | 0.004 | 34.0 | 0.007 |
| rs139141690 | G/A | 1.00 | 0.10 | 0.016 | 40.2 | 0.009 |
| rs342293 | C/G | 0.54 | -0.03 | 0.002 | 269.8 | 0.059 |
| rs74750282 | T/C | 0.91 | -0.05 | 0.004 | 164.1 | 0.036 |
| rs602616 | C/G | 0.91 | 0.02 | 0.004 | 27.2 | 0.006 |
| rs10085457 | G/A | 0.67 | -0.01 | 0.002 | 16.9 | 0.004 |
| rs3110823 | A/C | 0.83 | -0.03 | 0.003 | 139.1 | 0.031 |
| rs7800558 | T/C | 0.58 | 0.01 | 0.002 | 33.2 | 0.007 |
| rs117728810 | G/A | 0.94 | 0.02 | 0.005 | 27.4 | 0.006 |
| 8:6701534 | ACTC/A | 0.50 | -0.01 | 0.002 | 31.4 | 0.007 |
| rs4284061 | T/A | 0.56 | 0.02 | 0.002 | 84.2 | 0.020 |
| rs4841132 | A/G | 0.09 | 0.03 | 0.004 | 57.2 | 0.013 |
| 8:103218144 | ATTGCTATTATAAATAAGCTT/A | 0.94 | -0.03 | 0.004 | 44.0 | 0.010 |
| rs6986601 | A/G | 0.46 | 0.01 | 0.002 | 32.1 | 0.007 |
| rs385893 | T/C | 0.48 | -0.01 | 0.002 | 46.5 | 0.010 |
| 9:4838587 | TAC/T | 0.11 | -0.02 | 0.003 | 39.2 | 0.009 |
| rs7033052 | G/C | 0.47 | 0.01 | 0.002 | 18.2 | 0.004 |
| rs12247015 | A/G | 0.58 | -0.04 | 0.002 | 393.4 | 0.087 |
| rs7896518 | A/G | 0.57 | 0.05 | 0.002 | 578.1 | 0.130 |
| rs73349121 | G/C | 0.98 | 0.13 | 0.008 | 274.7 | 0.060 |
| rs7902510 | C/T | 0.79 | 0.03 | 0.003 | 153.3 | 0.034 |
| rs11594179 | C/T | 0.76 | -0.02 | 0.002 | 78.9 | 0.017 |
| rs7080536 | G/A | 0.96 | 0.03 | 0.005 | 38.2 | 0.009 |
| rs4910886 | G/T | 0.66 | -0.02 | 0.002 | 103.7 | 0.023 |
| rs2241942 | G/A | 0.76 | 0.01 | 0.002 | 31.6 | 0.007 |
| rs11235573 | T/C | 0.45 | 0.01 | 0.002 | 28.3 | 0.006 |
| rs74472890 | T/C | 0.95 | 0.03 | 0.005 | 30.2 | 0.007 |
| rs1362214 | A/G | 0.52 | -0.02 | 0.002 | 131.2 | 0.029 |
| rs1127787 | G/A | 0.83 | 0.02 | 0.003 | 45.2 | 0.010 |
| rs2015599 | G/A | 0.54 | -0.01 | 0.002 | 32.9 | 0.007 |
| rs6580981 | G/A | 0.54 | -0.01 | 0.002 | 45.3 | 0.010 |
| rs1716505 | C/G | 0.68 | 0.01 | 0.002 | 33.8 | 0.008 |
| rs12426673 | G/T | 0.42 | 0.01 | 0.002 | 45.9 | 0.010 |
| rs749140768 | AGGCACCTCTTCACAGGAC/A | 0.92 | -0.03 | 0.004 | 54.9 | 0.012 |
| rs11553699 | A/G | 0.86 | -0.05 | 0.003 | 252.5 | 0.060 |
| rs7987027 | T/C | 0.46 | -0.01 | 0.002 | 27.9 | 0.006 |
| rs1760940 | A/C | 0.75 | -0.03 | 0.002 | 124.4 | 0.027 |
| rs2771358 | T/C | 0.75 | 0.01 | 0.002 | 24.9 | 0.005 |
| rs17477725 | C/G | 0.41 | 0.01 | 0.002 | 20.5 | 0.005 |
| rs4427713 | T/C | 0.45 | 0.01 | 0.002 | 32.0 | 0.007 |
| rs117948349 | G/A | 0.97 | 0.03 | 0.006 | 34.7 | 0.008 |
| rs59488041 | T/A | 0.86 | 0.02 | 0.003 | 60.1 | 0.013 |
| rs261290 | T/C | 0.35 | -0.01 | 0.002 | 32.4 | 0.007 |
| rs141227171 | G/C | 1.00 | 0.09 | 0.023 | 15.8 | 0.004 |
| rs3087374 | C/A | 0.92 | -0.02 | 0.004 | 38.0 | 0.008 |
| rs151234 | G/C | 0.87 | 0.02 | 0.003 | 41.5 | 0.009 |
| rs289713 | T/A | 0.19 | 0.02 | 0.003 | 48.5 | 0.011 |
| rs55823018 | C/T | 0.68 | 0.01 | 0.002 | 33.8 | 0.008 |
| rs7213347 | G/C | 0.30 | 0.01 | 0.002 | 33.2 | 0.007 |
| rs12451698 | A/G | 0.76 | 0.02 | 0.002 | 62.7 | 0.014 |
| rs12601687 | G/A | 0.89 | -0.03 | 0.003 | 80.1 | 0.018 |
| rs1967556 | T/G | 0.47 | -0.02 | 0.002 | 117.8 | 0.026 |
| rs17850455 | C/G | 0.99 | -0.12 | 0.010 | 138.3 | 0.035 |
| rs11867543 | C/T | 0.86 | -0.02 | 0.003 | 36.7 | 0.008 |
| rs680478 | C/T | 0.25 | 0.02 | 0.002 | 46.0 | 0.010 |
| rs77261872 | C/T | 0.87 | -0.03 | 0.003 | 98.4 | 0.022 |
| rs17758695 | C/T | 0.97 | 0.04 | 0.006 | 47.1 | 0.010 |
| rs28665408 | A/C | 0.43 | -0.02 | 0.002 | 91.1 | 0.020 |
| rs12955015 | C/A | 0.02 | 0.04 | 0.007 | 30.6 | 0.007 |
| rs10411696 | T/G | 0.53 | 0.01 | 0.002 | 30.9 | 0.007 |
| rs11085147 | C/T | 0.90 | -0.09 | 0.004 | 634.4 | 0.147 |
| rs3218221 | G/A | 1.00 | 0.10 | 0.017 | 33.5 | 0.008 |
| rs142158911 | G/A | 0.88 | -0.02 | 0.003 | 28.1 | 0.006 |
| rs57843631 | C/T | 0.98 | -0.06 | 0.008 | 50.2 | 0.012 |
| rs139891465 | C/T | 0.96 | -0.05 | 0.005 | 85.7 | 0.019 |
| rs10419397 | G/A | 0.71 | 0.03 | 0.002 | 196.7 | 0.043 |
| rs35586766 | G/A | 0.91 | -0.04 | 0.004 | 109.3 | 0.024 |
| 19:19756073 | AGCC/A | 0.93 | -0.02 | 0.004 | 32.2 | 0.007 |
| rs7412 | C/T | 0.92 | -0.04 | 0.004 | 108.0 | 0.024 |
| rs11667430 | A/G | 0.60 | -0.01 | 0.002 | 15.6 | 0.004 |
| rs1613662 | G/A | 0.17 | 0.02 | 0.003 | 50.0 | 0.011 |
| rs11668201 | A/T | 0.81 | 0.02 | 0.003 | 31.3 | 0.007 |
| rs11696739 | G/A | 0.62 | 0.02 | 0.002 | 76.7 | 0.017 |
| rs156355 | T/C | 0.54 | -0.02 | 0.002 | 128.5 | 0.030 |
| rs4814776 | C/A | 0.67 | 0.04 | 0.002 | 262.4 | 0.058 |
| rs185387034 | A/G | 0.99 | 0.06 | 0.012 | 29.2 | 0.007 |
| rs754169 | T/A | 0.46 | -0.03 | 0.002 | 169.6 | 0.037 |
| rs76599088 | C/T | 0.98 | -0.08 | 0.008 | 94.1 | 0.021 |
| rs2426092 | A/C | 0.55 | -0.01 | 0.002 | 34.3 | 0.008 |
| rs577050795 | C/CA | 0.52 | -0.01 | 0.002 | 26.7 | 0.006 |
| rs2245947 | G/T | 0.32 | -0.04 | 0.002 | 301.2 | 0.066 |
| rs75107793 | G/A | 0.93 | -0.03 | 0.004 | 54.6 | 0.012 |
| rs12148 | T/G | 0.39 | 0.02 | 0.002 | 59.9 | 0.013 |

SNP: single-nucleotide polymorphisms; EA/Non-EA: effect allele/ Non effect allele; EAF: effect allele frequency; PVE: proportional variance explained by each SNP.

### Table S9 Mendelian Randomization results of mtDNA copy number on the risk of CAD (Instruments retrieved from Longchamps *et al*.)

|  | **Estimated effect** | **Q (p value) for heterogeneity** | **MR-Egger intercept (p value)** | **No. of outliers in MR-PRESSO** | **P for MR-PRESSO global test** |
| --- | --- | --- | --- | --- | --- |
| **CARDIoGRAMplusC4D** |  |  |  |  |  |
| IVW | 1.19 (1.01, 1.39) | 243.5 (0.0) |  |  |  |
| WME | 1.13 (0.96, 1.34) |  |  |  |  |
| MR Egger | 1.09 (0.79, 1.50) | 242.6 (0.0) | -0.002 (0.5) |  |  |
| MR PRESSO (outlier-corrected) | 1.14 (1.02, 1.27) |  |  | 4 | <0.001 |
| **UK Biobank** |  |  |  |  |  |
| IVW | 1.14 (1.01, 1.27) | 348.6 (0.0) |  |  |  |
| WME | 1.01 (0.90, 1.12) |  |  |  |  |
| MR Egger | 1.01 (0.81, 1.26) | 343.7 (0.0) | -0.003 (0.2) |  |  |
| MR PRESSO (outlier-corrected) | 1.07 (0.99, 1.15) |  |  | 5 | <0.001 |
| **FinnGen Study** |  |  |  |  |  |
| IVW | 1.29 (1.04, 1.59) | 228.8 (0.0) |  |  |  |
| WME | 1.21 (0.96, 1.52) |  |  |  |  |
| MR Egger | 1.18 (0.80, 1.74) | 228.1 (0.0) | -0.003 (0.6) |  |  |
| MR PRESSO (outlier-corrected) | 1.16 (1.00, 1.35) |  |  | 3 | <0.001 |

CAD: coronary artery disease; CARDIoGRAMplusC4D: Coronary Artery Disease Genome-Wide Replication and Meta-analysis plus the Coronary Artery Disease Genetics. IVW: Inverse-variance weighting; WME: Weighted-Median estimator; MR PRESSO: Mendelian Randomization Pleiotropy RESidual Sum and Outlier.

### Table S10 Mendelian Randomization results mtDNA copy number on the risk of HF (Instruments retrieved retrieved from Longchamps *et al*.)

|  | **Estimated effect** | **Q (p value) for heterogeneity** | **MR-Egger intercept (p value)** | **No. of outliers in MR-PRESSO** | **P for MR-PRESSO global test** |
| --- | --- | --- | --- | --- | --- |
| **HERMES consortium** |  |  |  |  |  |
| IVW | 1.02 (0.93, 1.13) | 152.2 (0.0) |  |  |  |
| WME | 1.03 (0.91, 1.17) |  |  |  |  |
| MR Egger | 1.08 (0.88, 1.33) | 151.5 (0.0) | 0.002 (0.5) |  |  |
| MR PRESSO (outlier-corrected) | 1.04 (0.95, 1.14) |  |  | 1 | 0.001 |
| **FinnGen Study** |  |  |  |  |  |
| IVW | 1.02 (0.89, 1.16) | 122.5 (0.0) |  |  |  |
| WME | 1.06 (0.88, 1.27) |  |  |  |  |
| MR Egger | 1.09 (0.85, 1.39) | 122.0 (0.0) | 0.003 (0.5) |  |  |
| MR PRESSO (outlier-corrected) | NA |  |  | 0 | 1.0 |

HF: heart failure; HERMES consortium: Heart Failure Molecular Epidemiology for Therapeutic Targets consortium. IVW: Inverse-variance weighting; WME: Weighted-Median estimator; MR PRESSO: Mendelian Randomization Pleiotropy RESidual Sum and Outlier. NA: not available.

UK Biobank data of heart failure was already integrated into HERMES consortium.

### Table S11 Genetic instruments at genome-wide significant level for mtDNA copy numberin the sensitivity MR analysis (Instruments retrieved from Hägg *et al.*)

| **SNP** | **EA/Non-EA** | **EAF** | **Beta** | **se** | **p value** | **F statistics** | **PVE (%)** |
| --- | --- | --- | --- | --- | --- | --- | --- |
| rs4648452 | T/C | 0.19 | -0.019 | 0.003 | 5.96E-09 | 33.8 | 0.011 |
| rs1474868 | T/C | 0.47 | -0.020 | 0.003 | 3.96E-15 | 61.7 | 0.020 |
| rs831522 | C/A | 0.36 | 0.023 | 0.003 | 7.69E-17 | 69.5 | 0.024 |
| 3:39163813_ga_g | G/GA | 0.12 | 0.023 | 0.004 | 1.82E-08 | 31.7 | 0.011 |
| rs35734242 | C/T | 0.44 | -0.014 | 0.003 | 4.36E-08 | 30.0 | 0.010 |
| rs12500975 | C/T | 0.43 | -0.015 | 0.003 | 1.02E-08 | 32.8 | 0.011 |
| rs518867 | T/C | 0.36 | -0.025 | 0.003 | 1.99E-20 | 85.8 | 0.028 |
| rs2853672 | A/C | 0.49 | -0.018 | 0.003 | 6.55E-12 | 47.2 | 0.016 |
| 5:88175199_tta_t | T/TTA | 0.47 | 0.015 | 0.003 | 8.57E-09 | 33.1 | 0.011 |
| rs114694170 | C/T | 0.05 | 0.049 | 0.005 | 3.27E-19 | 80.3 | 0.025 |
| rs2057657 | G/A | 0.23 | -0.018 | 0.003 | 3.81E-09 | 34.7 | 0.011 |
| rs210143 | T/C | 0.27 | -0.035 | 0.003 | 3.13E-36 | 158.0 | 0.050 |
| rs9469525 | A/G | 0.24 | 0.024 | 0.003 | 1E-13 | 55.4 | 0.021 |
| rs41294856 | T/C | 0.09 | -0.025 | 0.004 | 6.2E-09 | 33.8 | 0.010 |
| rs4895441 | G/A | 0.27 | 0.031 | 0.003 | 8.51E-28 | 119.4 | 0.039 |
| rs16874653 | G/A | 0.33 | -0.015 | 0.003 | 4.48E-08 | 29.9 | 0.010 |
| rs6976396 | A/C | 0.18 | -0.021 | 0.004 | 2.25E-09 | 35.7 | 0.013 |
| rs12155038 | A/G | 0.44 | -0.015 | 0.003 | 1.59E-08 | 31.9 | 0.011 |
| rs8179 | T/C | 0.22 | -0.024 | 0.003 | 2.89E-14 | 57.8 | 0.020 |
| rs342293 | G/C | 0.44 | 0.022 | 0.003 | 1.13E-17 | 73.3 | 0.024 |
| rs77236693 | T/C | 0.08 | 0.030 | 0.004 | 6.21E-12 | 47.3 | 0.014 |
| rs3110823 | C/A | 0.15 | 0.029 | 0.003 | 1.93E-17 | 72.2 | 0.021 |
| rs4841132 | A/G | 0.07 | 0.026 | 0.004 | 2.87E-09 | 35.3 | 0.010 |
| rs10094039 | A/G | 0.37 | -0.017 | 0.003 | 4.06E-11 | 43.6 | 0.014 |
| rs385893 | T/C | 0.49 | -0.022 | 0.003 | 7.21E-17 | 69.6 | 0.023 |
| rs10974817 | A/G | 0.41 | -0.015 | 0.003 | 3.1E-08 | 30.6 | 0.010 |
| rs56225686 | A/T | 0.05 | -0.032 | 0.005 | 1.64E-09 | 36.4 | 0.010 |
| rs11006121 | T/C | 0.40 | 0.019 | 0.003 | 1.58E-12 | 50.0 | 0.017 |
| rs11006132 | G/A | 0.23 | 0.031 | 0.003 | 4.44E-24 | 102.5 | 0.034 |
| rs10740118 | C/G | 0.43 | -0.027 | 0.003 | 4.53E-25 | 107.0 | 0.036 |
| rs1408343 | G/A | 0.23 | -0.026 | 0.003 | 2.41E-19 | 80.9 | 0.024 |
| rs113422568 | A/G | 0.33 | 0.019 | 0.003 | 7.33E-12 | 46.9 | 0.016 |
| rs741735 | A/C | 0.24 | 0.017 | 0.003 | 1.49E-08 | 32.1 | 0.011 |
| rs10835226 | T/C | 0.32 | 0.018 | 0.003 | 4.9E-10 | 38.7 | 0.014 |
| rs35979828 | T/C | 0.09 | -0.034 | 0.005 | 1.98E-11 | 45.0 | 0.018 |
| rs4388979 | G/T | 0.38 | 0.019 | 0.003 | 3.2E-13 | 53.1 | 0.017 |
| rs3809272 | A/G | 0.31 | -0.023 | 0.003 | 6.61E-17 | 69.8 | 0.023 |
| rs11615667 | A/C | 0.09 | 0.027 | 0.004 | 4.76E-11 | 43.3 | 0.012 |
| rs1760940 | C/A | 0.25 | 0.028 | 0.003 | 3.55E-21 | 89.2 | 0.029 |
| rs59488041 | A/T | 0.15 | -0.032 | 0.004 | 1.18E-17 | 73.2 | 0.027 |
| rs3087374 | A/C | 0.09 | 0.027 | 0.005 | 1.08E-08 | 32.7 | 0.011 |
| rs11865642 | C/A | 0.18 | -0.018 | 0.003 | 3.06E-08 | 30.7 | 0.010 |
| rs12924138 | T/G | 0.39 | 0.015 | 0.003 | 1.24E-08 | 32.4 | 0.010 |
| rs2063185 | T/C | 0.31 | 0.017 | 0.003 | 6.2E-09 | 33.8 | 0.012 |
| rs144382588 | A/AATC | 0.08 | 0.025 | 0.005 | 4.78E-08 | 29.8 | 0.010 |
| rs12451555 | G/T | 0.24 | -0.017 | 0.003 | 1.98E-08 | 31.5 | 0.011 |
| rs55757004 | T/G | 0.09 | -0.032 | 0.004 | 8.3E-13 | 51.2 | 0.016 |
| rs11078935 | G/T | 0.36 | 0.031 | 0.003 | 1.1E-31 | 137.2 | 0.045 |
| rs12941465 | A/G | 0.26 | -0.020 | 0.003 | 2.8E-11 | 44.3 | 0.015 |
| rs16978036 | T/G | 0.15 | 0.026 | 0.004 | 1.06E-11 | 46.2 | 0.017 |
| rs1790961 | T/G | 0.48 | -0.018 | 0.003 | 2.1E-12 | 49.4 | 0.016 |
| rs8110045 | G/A | 0.30 | -0.015 | 0.003 | 2.15E-08 | 31.4 | 0.010 |
| rs4510145 | A/G | 0.45 | -0.017 | 0.003 | 1.25E-10 | 41.4 | 0.014 |
| rs806709 | A/G | 0.09 | 0.070 | 0.004 | 3.03E-62 | 277.4 | 0.077 |
| rs566777150 | AAAAAAAAAAAAAG/A | 0.45 | 0.023 | 0.003 | 1.44E-18 | 77.3 | 0.026 |
| rs10419397 | A/G | 0.26 | -0.028 | 0.003 | 7.4E-23 | 96.9 | 0.030 |
| rs35586766 | A/G | 0.11 | 0.036 | 0.004 | 3.38E-16 | 66.6 | 0.025 |
| rs1065853 | T/G | 0.06 | 0.038 | 0.005 | 1.64E-15 | 63.5 | 0.017 |
| rs1613662 | G/A | 0.15 | 0.024 | 0.003 | 1.96E-12 | 49.5 | 0.015 |
| rs11696739 | A/G | 0.38 | -0.017 | 0.003 | 5.57E-11 | 43.0 | 0.014 |
| rs156333 | A/G | 0.48 | 0.020 | 0.003 | 3.64E-15 | 61.9 | 0.021 |
| rs11697739 | T/C | 0.48 | -0.025 | 0.003 | 5.4E-23 | 97.5 | 0.032 |
| rs11704452 | G/T | 0.17 | -0.026 | 0.004 | 2.98E-13 | 53.2 | 0.019 |
| rs760699 | T/C | 0.13 | -0.025 | 0.004 | 7.36E-11 | 42.4 | 0.014 |
| rs75107793 | A/G | 0.06 | 0.044 | 0.005 | 4.66E-19 | 79.6 | 0.022 |
| rs113214100 | CA/C | 0.17 | -0.020 | 0.004 | 7.58E-09 | 33.4 | 0.012 |

SNP: single-nucleotide polymorphisms; EA/Non-EA: effect allele/ Non effect allele; EAF: effect allele frequency; PVE: proportional variance explained by each SNP.

### Table S12 Mendelian Randomization results of mtDNA copy number on the risk of CAD (Instruments retrieved from Hägg *et al.*)

|  | **Estimated effect** | **Q (p value) for heterogeneity** | **MR-Egger intercept (p value)** | **No. of outliers in MR-PRESSO** | **P for MR-PRESSO global test** |
| --- | --- | --- | --- | --- | --- |
| **CARDIoGRAMplusC4D** |  |  |  |  |  |
| IVW | 1.18 (1.02, 1.38) | 82.3 (0.02) |  |  |  |
| WME | 1.13 (0.93, 1.39) |  |  |  |  |
| MR Egger | 1.29 (0.78, 2.12) | 82.1 (0.01) | 0.002 (0.7) |  |  |
| MR PRESSO (outlier-corrected) | 1.23 (1.07,1.42) |  |  | 1 | 0.03 |
| **UK Biobank** |  |  |  |  |  |
| IVW | 1.16 (1.00, 1.36) | 237.4 (0.0) |  |  |  |
| WME | 1.09 (0.95, 1.24) |  |  |  |  |
| MR Egger | 1.14 (0.71, 1.84) | 237.3 (0.0) | 0.0 (0.94) |  |  |
| MR PRESSO (outlier-corrected) | 1.16 (1.04, 1.30) |  |  | 3 | < 0.001 |
| **FinnGen Study** |  |  |  |  |  |
| IVW | 1.07 (0.85, 1.35) | 101.8 (0.0) |  |  |  |
| WME | 1.08 (0.83, 1.39) |  |  |  |  |
| MR Egger | 1.29 (0.62, 2.68) | 101.2 (0.0) | 0.005 (0.6) |  |  |
| MR PRESSO (outlier-corrected) | 1.05 (0.87, 1.28) |  |  | 2 | 0.001 |

CAD: coronary artery disease; CARDIoGRAMplusC4D: Coronary Artery Disease Genome-Wide Replication and Meta-analysis plus the Coronary Artery Disease Genetics. IVW: Inverse-variance weighting; WME: Weighted-Median estimator; MR PRESSO: Mendelian Randomization Pleiotropy RESidual Sum and Outlier.

### Table S13 Mendelian Randomization results mtDNA copy number on the risk of HF (Instruments retrieved from Hägg *et al.*)

|  | **Estimated effect** | **Q (p value) for heterogeneity** | **MR-Egger intercept (p value)** | **No. of outliers in MR-PRESSO** | **P for MR-PRESSO global test** |
| --- | --- | --- | --- | --- | --- |
| **HERMES consortium** |  |  |  |  |  |
| IVW | 1.01 (0.89, 1.14) | 75.4 (0.05) |  |  |  |
| WME | 1.05 (0.90, 1.23) |  |  |  |  |
| MR Egger | 1.32 (0.91, 1.90) | 72.5 (0.07) | 0.007 (0.1) |  |  |
| MR PRESSO (outlier-corrected) | 1.05 (0.94,1.17) |  |  | 1 | 0.054 |
| **FinnGen Study** |  |  |  |  |  |
| IVW | 0.97 (0.80, 1.17) | 89.9 (0.0) |  |  |  |
| WME | 1.09 (0.87, 1.38) |  |  |  |  |
| MR Egger | 1.13 (0.63, 2.04) | 89.4 (0.0) | 0.004 (0.6) |  |  |
| MR PRESSO (outlier-corrected) | 1.04 (0.89, 1.22) |  |  | 1 | 0.004 |

HF: heart failure; HERMES consortium: Heart Failure Molecular Epidemiology for Therapeutic Targets consortium. IVW: Inverse-variance weighting; WME: Weighted-Median estimator; MR PRESSO: Mendelian Randomization Pleiotropy RESidual Sum and Outlier.

UK Biobank data of heart failure was already integrated into HERMES consortium.

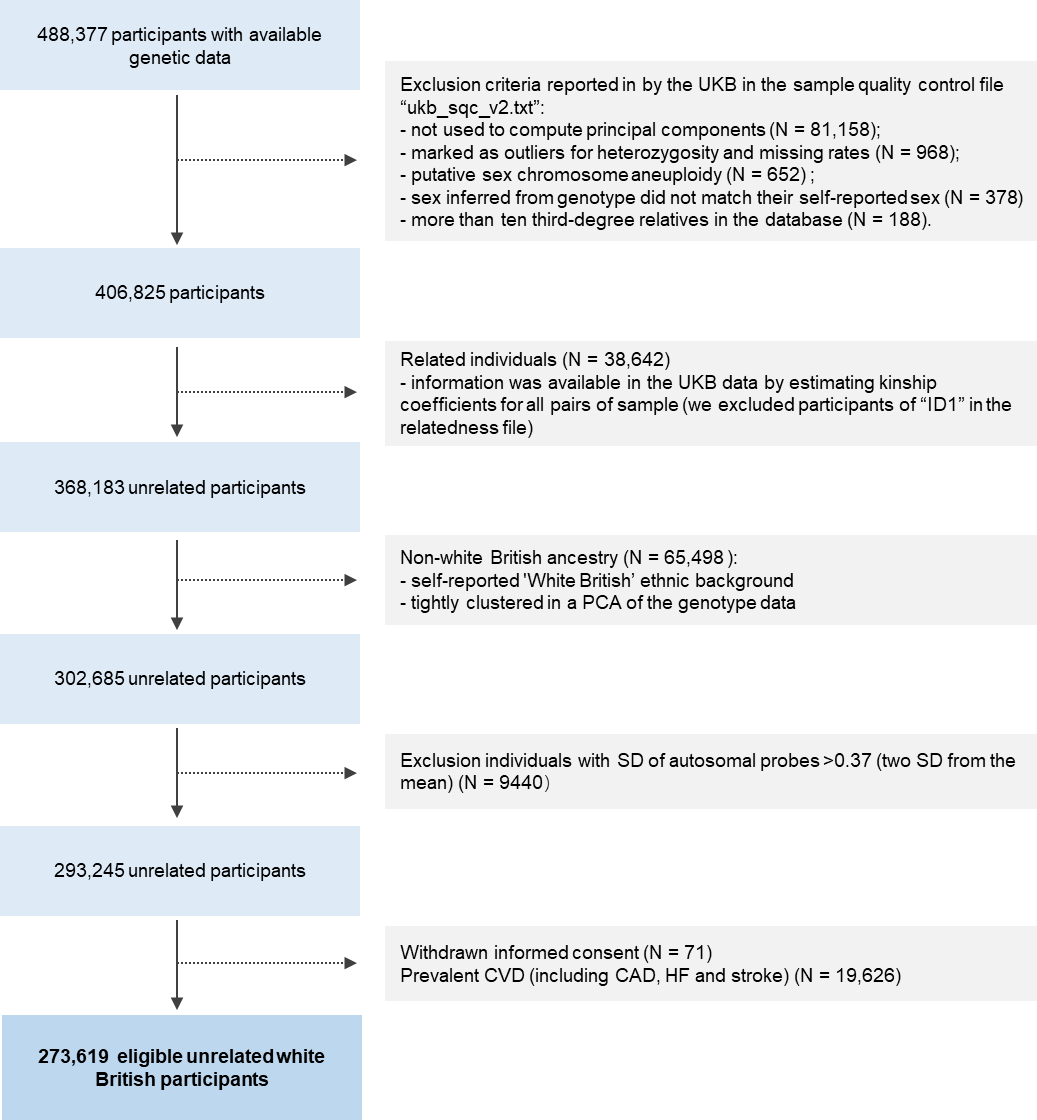

### Figure S1 Flowchart of participants inclusion in UK Biobank

PCA: Principal components analysis; CVD: cardiovascular disease; CAD: coronary artery disease; HF: heart failure.

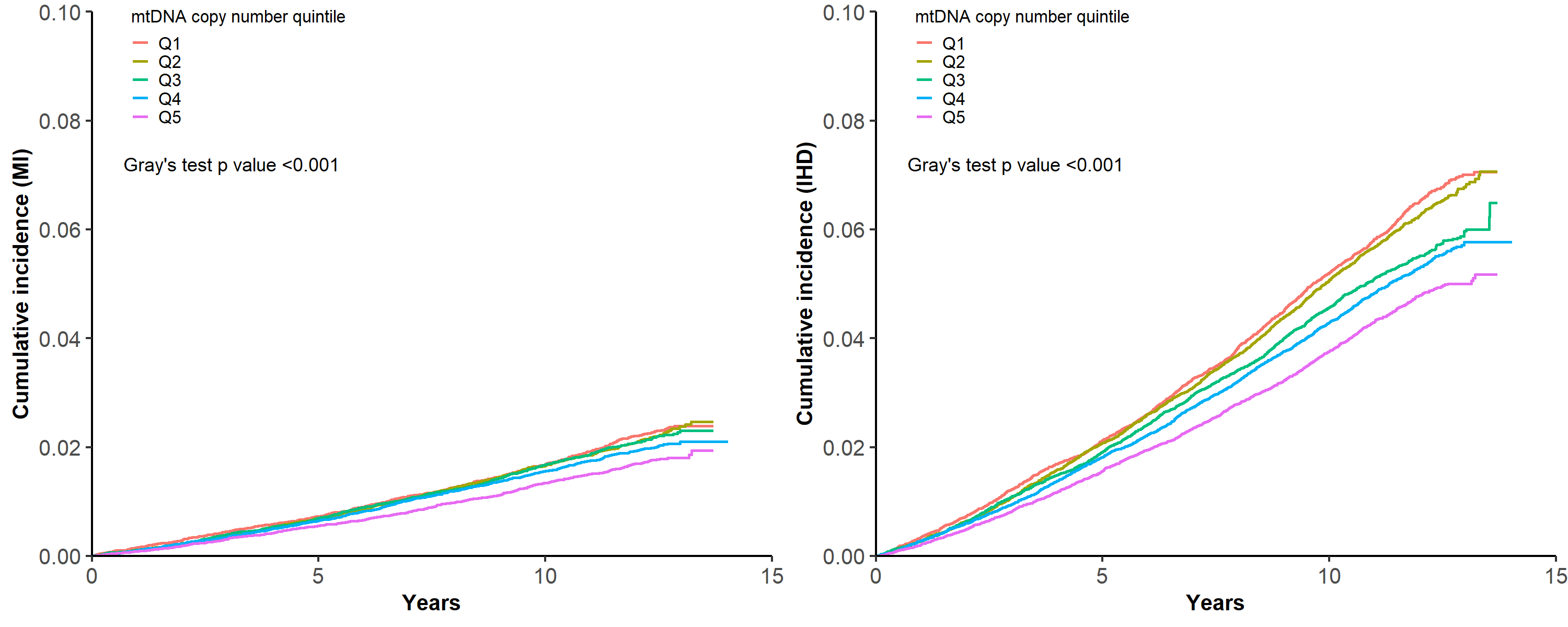

### Figure S2 Cumulative Incidence of MI and IHD by quintiles of mtDNA copy number

We calculated Cumulative incidence for CAD and HF, accounting for death as a competing event. Differences in cumulative incidence between mtDNA copy number quintiles were assessed using Gray’s test.

MI: myocardial infarction; IHD: ischemic heart disease.

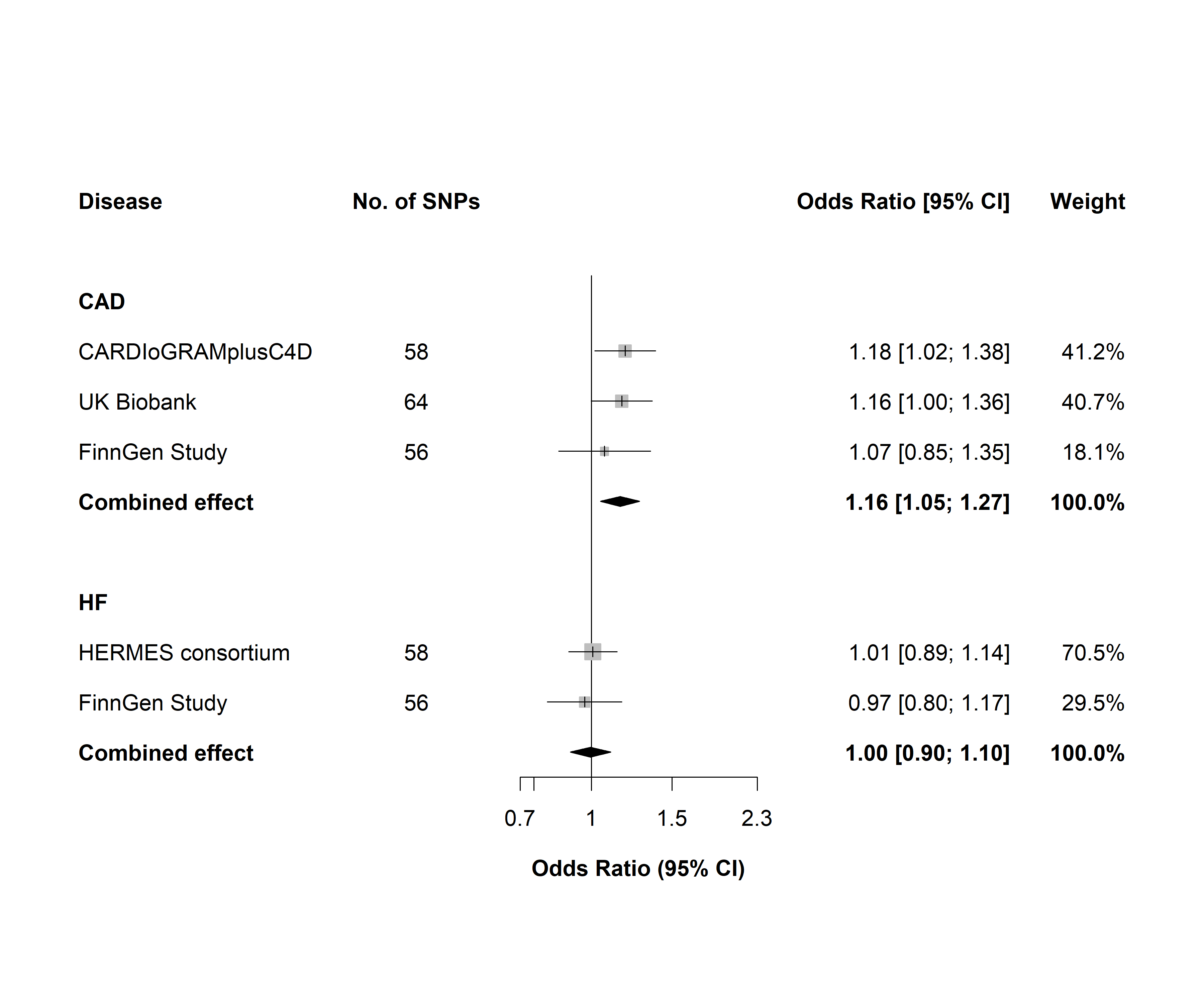

### Figure S3 Mendelian Randomization study of mtDNA copy number on the risk of CAD and HF (Instruments retrieved from Hägg et al.)

Estimated ORs for the effect of per-SD decrease in mtDNA copy number on the risk of CAD and HF, obtained from an MR inverse-variance weighted method, per outcome database separately and combined over the different databases using fixed-effect meta-analyses.

CAD: coronary artery disease; HF: heart failure; CARDIoGRAMplusC4D: Coronary Artery Disease Genome-Wide Replication and Meta-analysis plus the Coronary Artery Disease Genetics; HERMES consortium: Heart Failure Molecular Epidemiology for Therapeutic Targets consortium.

UK Biobank data of heart failure was already integrated into HERMES consortium.

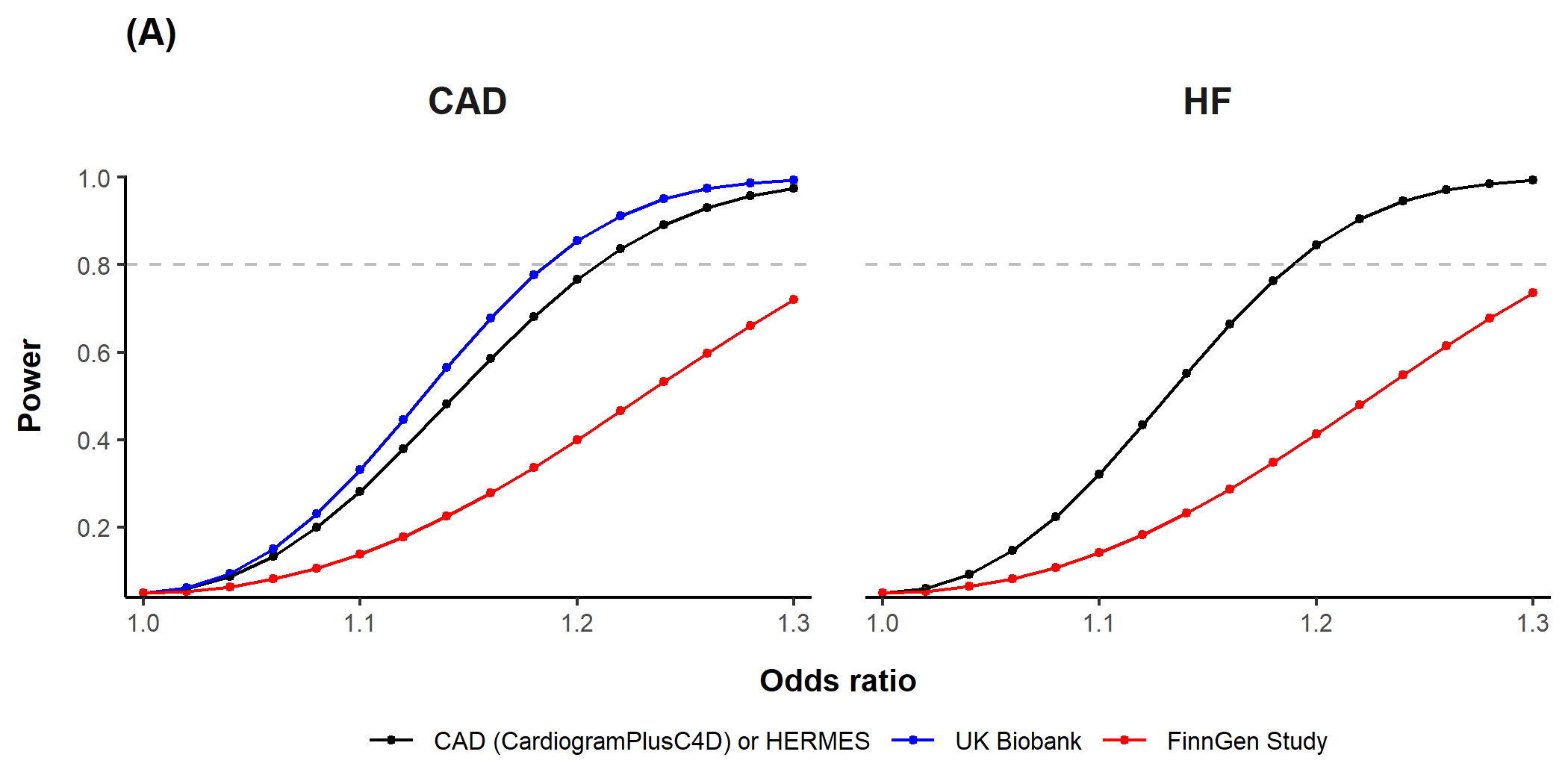

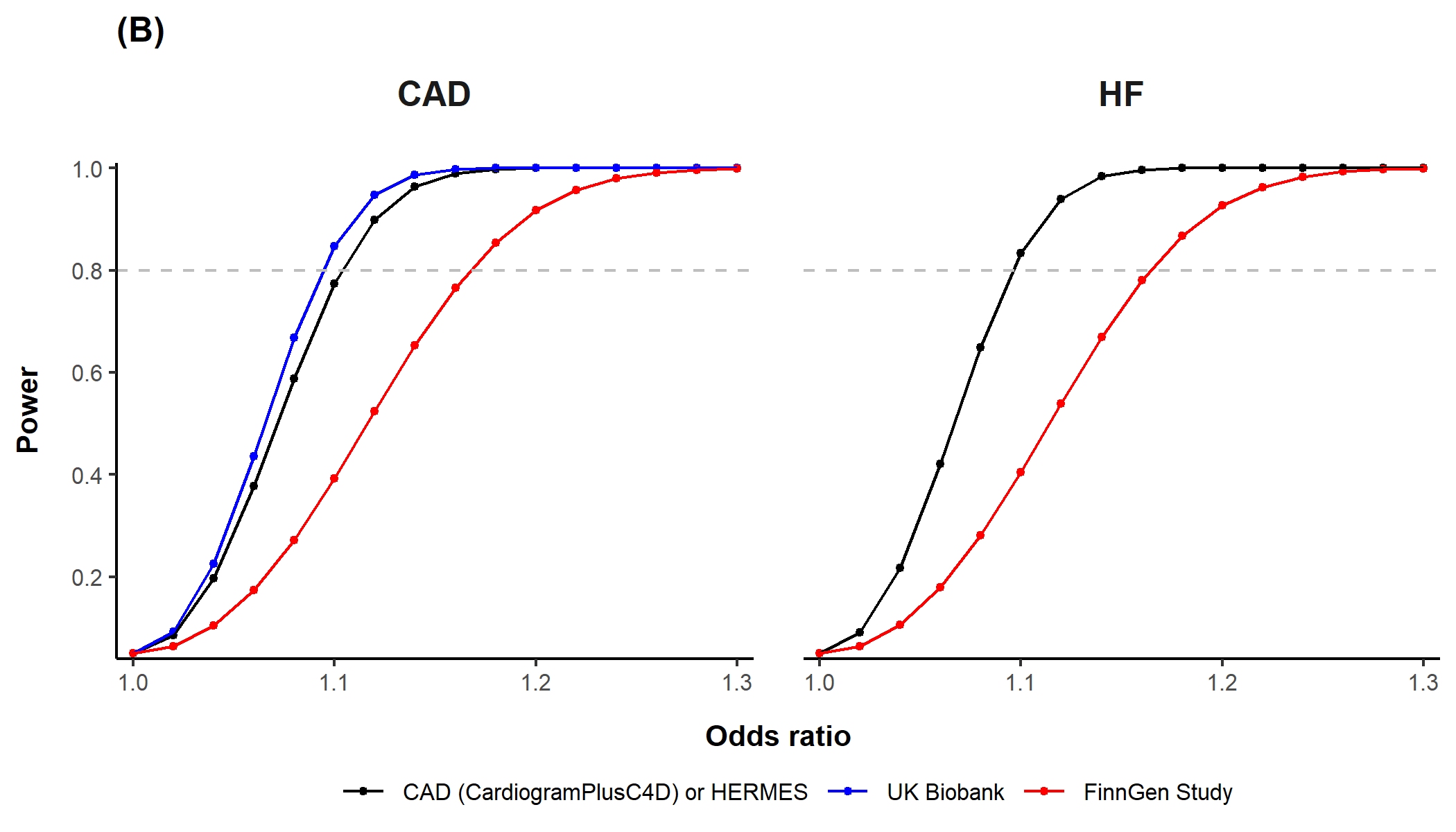

### Figure S4 Statistical power of Mendelian Randomization analyses

Statistical power was calculated via the online tool for binary outcomes (<https://github.com/kn3in/mRnd>) (4), where the alpha level was set to 0.05. Dashed grey line indicates statistical power of 0.8. (A) Power calculation for the main MR analyses using genetic variants identified by Longchamps *et al.*; though the calculated variation was 1.9%, total variation was set to 0.5%, which was argued in the paper that less than 1% of the variance in mtDNA copy number explained by GWAS loci when predicted into the Atherosclerosis Risk in Communities (ARIC) Study cohort. (B) Power calculation for the sensitivity MR analysis using genetic variants identified by Hägg *et al.*; total variation was set to 1.3% as calculated.
